# Post-Diarrheal Nutritional Trajectories Among Malnourished Children: A Clustering and Multinomial Modelling Approach

**DOI:** 10.64898/2026.04.20.26351264

**Authors:** Billy Ogwel, Alex O. Awuor, Brian O. Onyando, Raphael Ochieng, M. Jahangir Hossain, Bakary Conteh, Wardah Mujahid, Fariha Shaheen, Vitumbiko Munthali, Thandizo Malemia, Milagritos Tapia, Adama Mamby Keita, Dilruba Nasrin, Margaret N Kosek, Firdausi Qadri, Karen L. Kotloff, Patricia B. Pavlinac, Richard Omore, Elizabeth T Rogawski McQuade

**Author notes:** Correspondence: Billy Ogwel; KEMRI-CGHR, P.O Box 1578-40100, Kisumu, Kenya. These authors contributed equally to this work as joint senior authors (Richard Omore, Elizabeth T Rogawski McQuade).

## Abstract

Although the co-occurrence of diarrhea and malnutrition is well documented, research has largely focused on the acute management of diarrheal illness. Despite its importance, longitudinal evidence characterizing post-diarrheal recovery trajectories is sparse. We sought to characterize post-diarrheal nutritional recovery trajectories among children aged 6–35 months who were malnourished at enrollment using data from the Enterics for Global Health (EFGH) Shigella Surveillance study (2022–2024). EFGH enrolled children aged 6–35 months presenting with medically-attended diarrhea and followed them at 4 weeks and 3 months post-enrollment. This analysis included children with baseline wasting, stunting, or underweight (z-score < −2) and complete anthropometric follow-up. Latent class mixed-effects models were used to identify distinct post-diarrheal growth trajectories based on changes in anthropometric z-scores over time. Multinomial modified Poisson regression models examined associations between baseline factors and trajectory membership. Among 9,480 enrolled children, 16.5% (n=1,561) were wasted, 22.7% (n=2,155) stunted, and 21.0% (n=1,994) underweight at baseline. Wasting showed greater recovery potential (80.8%) compared with stunting (38.5%) and underweight (40.3%). Recovery was shaped by factors across multiple levels. Clinical severity markers ( prolonged diarrhea, dehydration, and hypoxemia) increased the risk of nutritional failure. Age also influenced outcomes: infants were more likely to worsen, whereas older toddlers more often experienced stagnation. Interventions including exclusive breastfeeding, oral rehydration therapy, appropriate antibiotics, and zinc supplementation, improved outcomes, while unimproved sanitation undermined recovery. These findings highlight the need for integrated strategies combining infection control, nutritional rehabilitation, and water, sanitation, and hygiene interventions tailored to the children’s developmental stage.

**Key Messages:**

- Post-diarrheal nutritional recovery is highly heterogeneous, with wasting showing the greatest potential for improvement, while stunting and underweight often result in persistent growth stagnation.
- Baseline anthropometric deficits alone are insufficient to predict recovery, highlighting the need for dynamic monitoring and individualized management.
- Infants are particularly vulnerable to acute nutritional deterioration, while older toddlers frequently experience growth stagnation.
- Modifiable protective factors including exclusive breastfeeding, ORS, zinc, and appropriate antibiotics, improved outcomes, whereas poor sanitation undermined recovery.
- Integrated strategies, tailored to a child’s developmental stage, combining clinical care, nutrition, and environmental interventions are critical to support sustained child growth and development.

## Introduction

Malnutrition contributes to approximately 50% of under-five deaths, predominantly occurring in low-and middle-income countries (LMICs) [1]. The bidirectional relationship between enteric infections and malnutrition remains one of the most formidable barriers to improving child survival and developmental potential in LMICs. Diarrhea itself is a primary etiological factor in childhood malnutrition in LMICs [2], driven by anorexia, impaired nutrient absorption, intestinal mucosal damage, and nutrient depletion. Moreover, the self-perpetuating cycle between diarrhea and malnutrition is extended as malnutrition compromises the body’s immune system, increasing susceptibility to future diarrheal episodes, leading to more frequent and prolonged illness. This vicious cycle in early childhood can also potentially predispose children to increased risk of cardiovascular disease, type 2 diabetes, and obesity in adulthood [3,4].

While the co-occurrence of diarrhea and malnutrition is well-documented [5,6], scientific and clinical focus has predominantly centered on acute management: rehydration, infection treatment, and nutritional stabilization. A critical, yet overlooked, phase is the post-diarrheal recovery period, physiological window that determines long-term developmental trajectories. For children already presenting with wasting, stunting, or underweight, each diarrheal episode represents a potential catalyst for further growth deceleration and metabolic reprogramming with lifelong consequences. Most existing evidence characterizes post-diarrheal growth using average changes in anthropometric indices, implicitly assuming homogeneous recovery patterns [7].

Wasted, stunted, and underweight children may follow fundamentally different recovery pathways after diarrhea due to differences in physiological reserves, chronicity of malnutrition, age-related growth potential, and exposure to adverse social and environmental conditions. Disaggregating these trajectories is therefore essential for identifying high-risk children, refining prognostic assessment, and tailoring post-diarrheal care strategies in high-burden settings. Moreover, diarrheal episodes represent a critical opportunity for intervention. Health system contact during illness potentiates improving trajectories through timely case management, nutritional support, and targeted follow-up.

Despite its importance, longitudinal evidence characterizing post-diarrheal recovery trajectories is sparse. In this context, we aimed to characterize post-diarrheal nutritional trajectories among children aged 6-35 months who were wasted, stunted, or underweight at enrollment using longitudinal anthropometric data from the Enterics for Global Health (EFGH) Shigella Surveillance study [8].

## Methods

### Study Design and Population

We conducted a secondary analysis of the EFGH Shigella surveillance study, a two-year prospective study conducted at 7 country sites (Bangladesh, Kenya, Malawi, Mali, Pakistan, Peru and The Gambia) [9–15] using cross-sectional and longitudinal study designs between 2022–2024. EFGH’s primary objective was to characterize the epidemiology of Shigella attributable MAD, including incidence and sequelae, utilizing both cross-sectional and longitudinal observational designs. The study design, epidemiological and data procedures, and laboratory methods for EFGH are described in detail elsewhere [8,16–18]. In summary, EFGH enrolled MAD cases defined as children aged 6-35 months presenting with diarrhea (≥ 3 looser-than-normal stools within 24 hours) that began within the past 7 days after ≥2 diarrhea-free days seeking care at recruitment centers in the study catchment area [8]. Follow-ups were conducted at week-4 (24-67 days) and month-3 (84-127 days) after enrolment.

We restricted this current analysis to EFGH participants who were either wasted, stunted or underweight at baseline (Z-scores less than −2) and had complete follow-up data at both week-4 and month-3.

#### Procedures

Anthropometric data (length[<24 months]/height[≥24 months], weight, and MUAC) were collected at enrollment, week 4, and month 3 using standardized equipment (ShorrBoard®, digital scales, and 25-cm tapes). Weight and mid-upper arm circumference (MUAC) were recorded post-rehydration, using tared weighing for children <24 months or those unable to stand. Measurements were performed in duplicate and averaged; a third reading was required if discrepancies exceeded predefined thresholds (weight ≥0.2 kg, MUAC ≥0.3 cm, length/height ≥0.5 cm). Rigorous quality control included weekly scale calibrations and semi-annual standardization tests against a site-specific gold standard measurer. Furthermore, 5% of all participants underwent remeasurement by the gold standard measurer to ensure sustained data accuracy [8].

Standardized data collection procedures, including socio-demographic information, epidemiological factors, illness history, clinical management details, and outcome measures, were implemented by trained study staff at each visit. In addition, a focused physical examination was performed.

#### Data preprocessing

Prior to analysis, the dataset underwent rigorous data quality control. Z-scores were calculated relative to the 2006 WHO Child Growth Standards [19]. Children exhibiting physiologically implausible anthropometric measurements, specifically height-for-age z-score (HAZ) outside the range of -6 to +6 SD, weight-for-height z-score (WHZ) outside the range of -5 to +5 SD, or weight-for-age Z-scores (WAZ) outside the range of -6 to +5 SD, were excluded [20].

Wasting was defined as a WHZ < -2 or a MUAC < 12.5 cm; stunting, as a HAZ < -2; and underweight, as a WAZ < -2.

#### Statistical Analysis

##### Latent Class Mixed-effects Modelling

The follow-up day variable was derived as the number of days between the enrolment date and the date of the follow-up visit. To ensure comparability of trajectories across children, baseline anthropometric values (WHZ, HAZ, WAZ) were included in the models as covariates. The outcome variable was defined as the change in the anthropometric score from enrolment (Δ-score = follow-up value − baseline value), such that each child’s trajectory represents deviation from their baseline measurement. Age categories of the children at enrolment, and sex, household wealth quintile, modified Vesikari score (MVS) classification, and antibiotic use at enrolment were included as covariates in the fixed effects part of the models.

Latent class mixed models (LCMM) were used to identify subgroups of children with distinct growth trajectories [21]. Models were estimated using the hlme() function from the lcmm package in R. For each outcome (wasting, stunting, and underweight), we compared solutions with 1 to 5 latent classes. The fixed-effects portion of the model specified Δ-score as a function of follow-up time (in days), baseline anthropometry, and child- and household-level covariates. Random intercepts at the individual level were included to account for within-child correlation. Class-specific trajectories were modeled through a mixture term that allowed the slope for follow-up day to vary across classes.

For each outcome, the one-class model was fit first, followed by models with 2–5 classes using a grid search procedure with multiple random starts (5 repetitions, maximum 30 iterations) to avoid convergence to local maxima. The optimal number of classes was selected by comparing log-likelihood, Akaike Information Criterion (AIC), Bayesian Information Criterion (BIC), and entropy. BIC was prioritized for class enumeration, with entropy used to assess classification quality [22]. Based on the above criteria, we identified the optimal number of classes to be four for wasting and stunting, and five for Underweight (Table S1).

##### Descriptive and Multinomial Modelling

Similar latent trajectories were collapsed into three clinically meaningful categories (Improving: positive growth slope; Worsening: negative growth slope; Stagnating: slope centered around zero) to mitigate small class sizes and enhance clinical interpretability during downstream modeling.

Analyses were conducted separately for wasting, stunting, and underweight cohorts. Baseline characteristics were summarized overall and stratified by nutritional trajectory class. Categorical variables were described using frequencies and column percentages, while continuous variables were summarized using medians and interquartile ranges (Q1–Q3). Results were presented in trajectory-stratified summary tables. Individual-level clinical, demographic, socioeconomic, and water, sanitation and hygiene data were evaluated as potential covariates. Associations between baseline factors and nutritional trajectory membership were evaluated using multinomial modified Poisson regression models, implemented via vector generalized linear models [23]. Separate models were fitted for wasting, stunting, and underweight outcomes. Nutritional trajectory class was treated as a nominal outcome with Improving as the base (reference) category. Results were expressed as risk ratios (RRs) with corresponding 95% confidence intervals (CIs). Each candidate predictor was first evaluated in a bivariate multinomial model. Predictors with a minimum p-value < 0.20 across outcome comparisons were retained for multivariable modeling. Predictors with no within-sample variation were excluded. Multivariable multinomial models included predictors passing bivariate screening, after excluding collinear variables identified a priori and through the Cramer’s V statistic. Adjusted risk ratios (aRRs) and 95% CIs were derived from exponentiated model coefficients. Statistical significance was assessed at a two-sided α = 0.05. Adjusted associations were visualized using forest plots, stratified by Worsening and Stagnating trajectories relative to the Improving reference group. Statistically significant associations were highlighted graphically.

All the statistical analyses were carried out using R software, version 4.5.2 (R Foundation for Statistical Computing, Vienna, Austria).

#### Ethical Considerations

The current study leveraged data from the EFGH study, which received ethical approvals from the Institutional Review Boards (IRBs) of all participating institutions, as detailed elsewhere [8]. Written informed consent was obtained from the caregivers of all children prior to any research procedures.

## Results

### Cluster Identification of Nutritional Trajectories for Growth Outcomes

During the study period a total of 9,480 children were recruited across the seven EFGH Country sites, of whom 16.5% (n=1,561), 22.7% (n=2,155) and 21.0% (n=1,994) were wasted , stunted and underweight, respectively (Figure 1). Latent class mixed-effects modelling identified four distinct wasting (WHZ) recovery trajectories following MAD episodes. The class-specific intercepts represent the model-estimated starting points of each WHZ trajectory at baseline (expected value at time zero) rather than the observed enrolment WHZ values. The largest group, Class 3 (n = 1,247), showed a moderately negative trajectory baseline (intercept = −0.26; p = 0.006) and a moderate positive recovery slope over follow-up (0.014 WHZ units/day; p < 0.001). Class 1 (n = 258) had a similar estimated starting point (−0.34; p < 0.001) but demonstrated a slower rate of improvement (0.004/day; p < 0.001). A small subgroup, Class 4 (n = 14), had the lowest estimated trajectory baseline (−1.74; p < 0.001) followed by a rapid recovery trajectory (0.045/day; p < 0.001). In contrast, Class 2 (n = 42) had the highest estimated trajectory baseline (0.45; p = 0.074)but experienced a progressive decline in nutritional status over time (−0.015/day; p < 0.001) (Table 1).

**Figure 1.**
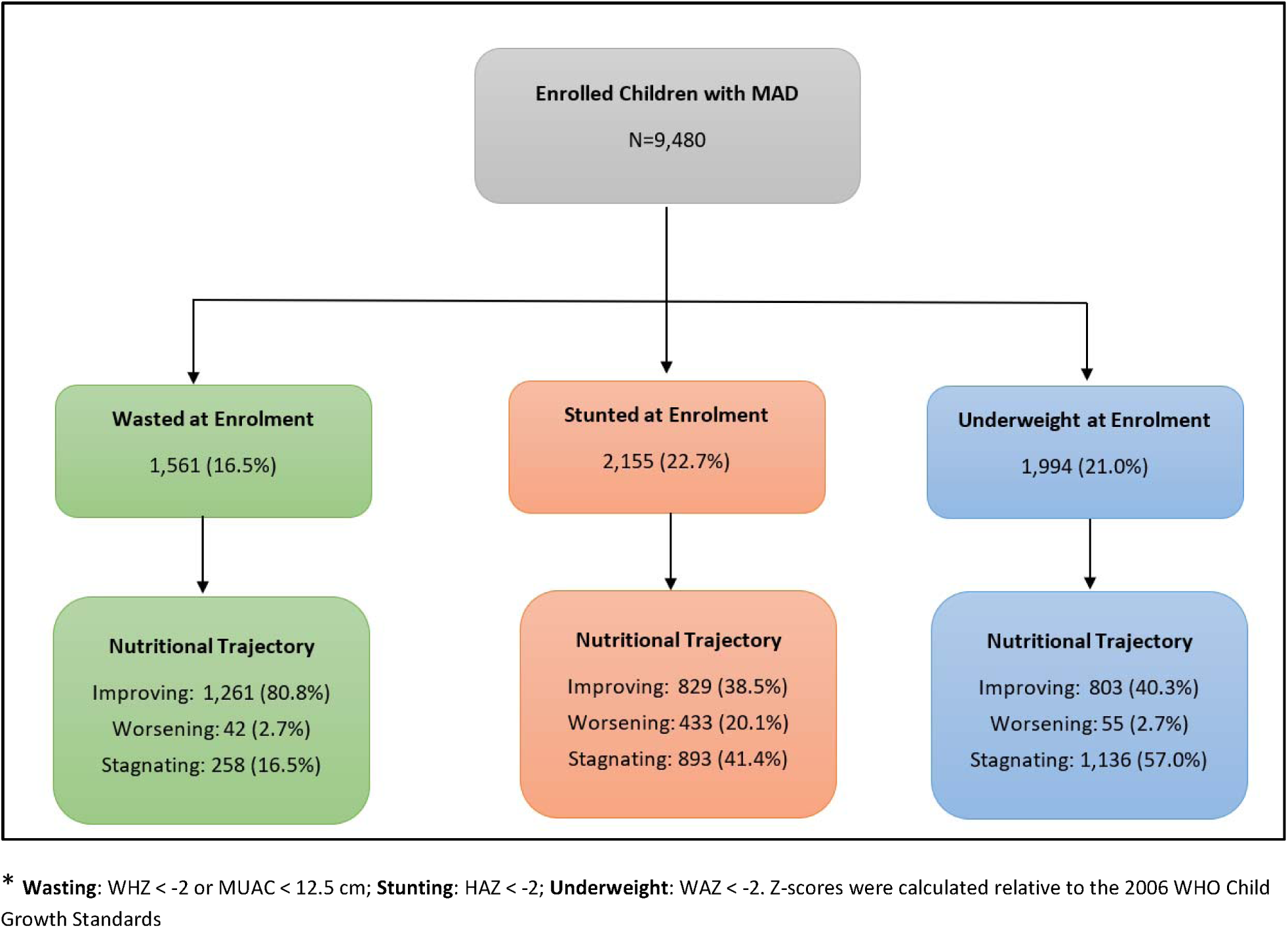
Enrolment flowchart and nutritional trajectories among malnourished children aged 6-35 months presenting with medically-attended diarrhea in the EFGH Study, 2022-2024

**Table 1.** Latent Class Mixed-Effects Model–Derived Nutritional Trajectories for Wasting (WHZ), Stunting (HAZ), and Underweight (WAZ) Following Medically Attended Diarrhea Episode.

| Nutritional Outcome |  | Coefficients | Standard Error | Wald Statistic | P-value |
| --- | --- | --- | --- | --- | --- |
| Wasting (N= 1,5461) | **Intercept** |  |  |  |  |
|  | intercept class1 (n=258) | -0.336 | 0.067 | -5.026 | <0.001 |
|  | intercept class2 (n= 42) | 0.452 | 0.254 | 1.785 | 0.074 |
|  | intercept class3 (n=1,247) | -0.260 | 0.094 | -2.772 | 0.006 |
|  | intercept class4 (n=14) | -1.739 | 0.371 | -4.688 | <0.001 |
|  | **Slope** |  |  |  |  |
|  | Follow-up days class1 | 0.004 | 0.001 | 7.449 | <0.001 |
|  | Follow-up days class2 | -0.015 | 0.002 | -8.794 | <0.001 |
|  | Follow-up days class3 | 0.014 | 0.001 | 10.827 | <0.001 |
|  | Follow-up days class4 | 0.045 | 0.004 | 12.782 | <0.001 |
| Stunting (N=2,155) | **Intercept** |  |  |  |  |
|  | intercept class1 (n=829) | -0.125 | 0.043 | -2.918 | 0.004 |
|  | intercept class2 (n=300) | -0.137 | 0.022 | -6.132 | <0.001 |
|  | intercept class3 (n=893) | -0.065 | 0.024 | -2.702 | 0.007 |
|  | intercept class4 (n=133) | 0.059 | 0.027 | 2.167 | 0.030 |
|  | **Slope** |  |  |  |  |
|  | Follow-up days class1 | 0.006 | 0.001 | 12.218 | <0.001 |
|  | Follow-up days class2 | -0.003 | 0.000 | -9.460 | <0.001 |
|  | Follow-up days class3 | 0.000 | 0.000 | 1.157 | 0.247 |
|  | Follow-up days class4 | -0.009 | 0.000 | -29.974 | <0.001 |
| Underweight (N=1,994) | **Intercept** |  |  |  |  |
|  | intercept class1 (n=471) | -0.073 | 0.040 | -1.811 | 0.070 |
|  | intercept class2 (n=312) | -0.518 | 0.164 | -3.165 | 0.002 |
|  | intercept class3 (n=20) | 0.135 | 0.075 | 1.810 | 0.070 |
|  | intercept class4 (n= 1,136) | -0.247 | 0.070 | -3.510 | 0.000 |
|  | intercept class5 (n= 55) | 0.948 | 0.296 | 3.209 | 0.001 |
|  | **Slope** |  |  |  |  |
|  | Follow-up days class1 | 0.002 | 0.001 | 3.368 | 0.001 |
|  | Follow-up days class2 | 0.019 | 0.002 | 9.108 | 0.000 |
|  | Follow-up days class3 | 0.006 | 0.001 | 5.554 | 0.000 |
|  | Follow-up days class4 | 0.000 | 0.001 | -0.755 | 0.450 |
|  | Follow-up days class5 | -0.012 | 0.002 | -6.933 | 0.000 |

**Wasting:** WHZ < -2 or MUAC < 12.5 cm; **Stunting:** HAZ < -2; **Underweight:** WAZ < -2
\* Respective baseline anthropometric values Age, sex, wealth quintile, modified Vesikari score, and antibiotic use included in LCMM models

Similarly, for stunting (HAZ) LCMM identified four distinct linear growth trajectories following MAD episode. The largest groups were Class 3 (n = 893) and Class 1 (n = 829), which had slightly negative estimated trajectory baselines (intercepts = −0.07 and −0.13, respectively; both p < 0.01). Class 2 (n = 300) had the lowest estimated trajectory baseline (−0.14; p < 0.001), whereas Class 4 (n = 133) had a slightly positive estimated baseline (0.06; p = 0.030). Over follow-up, Class 1 demonstrated modest linear growth recovery (0.006 HAZ units/day; p < 0.001). In contrast, Class 2 experienced progressive linear growth faltering (−0.003/day; p < 0.001), and Class 4 showed a marked decline in HAZ over time (−0.009/day; p < 0.001). Class 3 exhibited no significant change in linear growth during follow-up (slope ≈ 0.000; p = 0.247) (Table 1).

Among underweight children, LCMM identified five distinct post-diarrheal growth trajectories. At enrolment, the largest group, Class 4 (n = 1,136), had a moderately negative estimated trajectory baseline (WAZ = −0.25; p < 0.001) with no significant change over follow-up (slope ≈ 0; p = 0.450), indicating growth stagnation. Class 2 (n = 312) had the lowest estimated trajectory baseline (−0.52; p = 0.002) but demonstrated substantial recovery over time (0.019 WAZ units/day; p < 0.001). Class 1 (n = 471) and Class 3 (n = 20) had trajectory baselines close to the reference median (−0.07 and 0.14, respectively; both p = 0.070) and experienced modest improvements during follow-up (0.002/day and 0.006/day; both p ≤ 0.001). In contrast, Class 5 (n = 55) had the highest estimated trajectory baseline (0.95; p = 0.001) but underwent a significant decline in nutritional status over time (−0.012/day; p < 0.001) (Table 1).

### Cohort Characteristics and Nutritional Trajectories

Across the three nutritional cohorts, wasted (N=1,561), stunted (N=2,155), and underweight (N=1,994), latent trajectories followed distinct patterns. Improvement was most prevalent in the wasting cohort (80.8%), compared to only 38.5% of stunted and 40.3% of underweight children. Conversely, stagnation was the dominant trajectory for stunted (41.4%) and underweight (57.0%) children, while only 16.5% of wasted children stagnated. Worsening trajectories remained rare across all groups, ranging from 2.7% to 20.1% (Figure 1).

Across all three cohorts, children exhibited broadly similar demographic, socioeconomic, and clinical profiles. Median age ranged from 14 to 18 months, and the majority were male (53.2%–61.8%). Socio-economic vulnerability was pronounced across all cohorts, with 76.4%–83.2% of caregivers unemployed and fewer than 6% of households in the highest wealth quintile. Clinical severity was also comparable: approximately 21.3%–25.3% of episodes met criteria for severe diarrhea based on the Modified Vesikari score, 19.0%–21.2% of children presented with some dehydration, and ∼1.3–1.5% had severe dehydration. Across cohorts, children experienced a median of 6 loose stools (IQR: 4–8) with a median diarrhea duration of 3 days (IQR: 2–4) (Table 2,3 and 4).

**Table 2:**
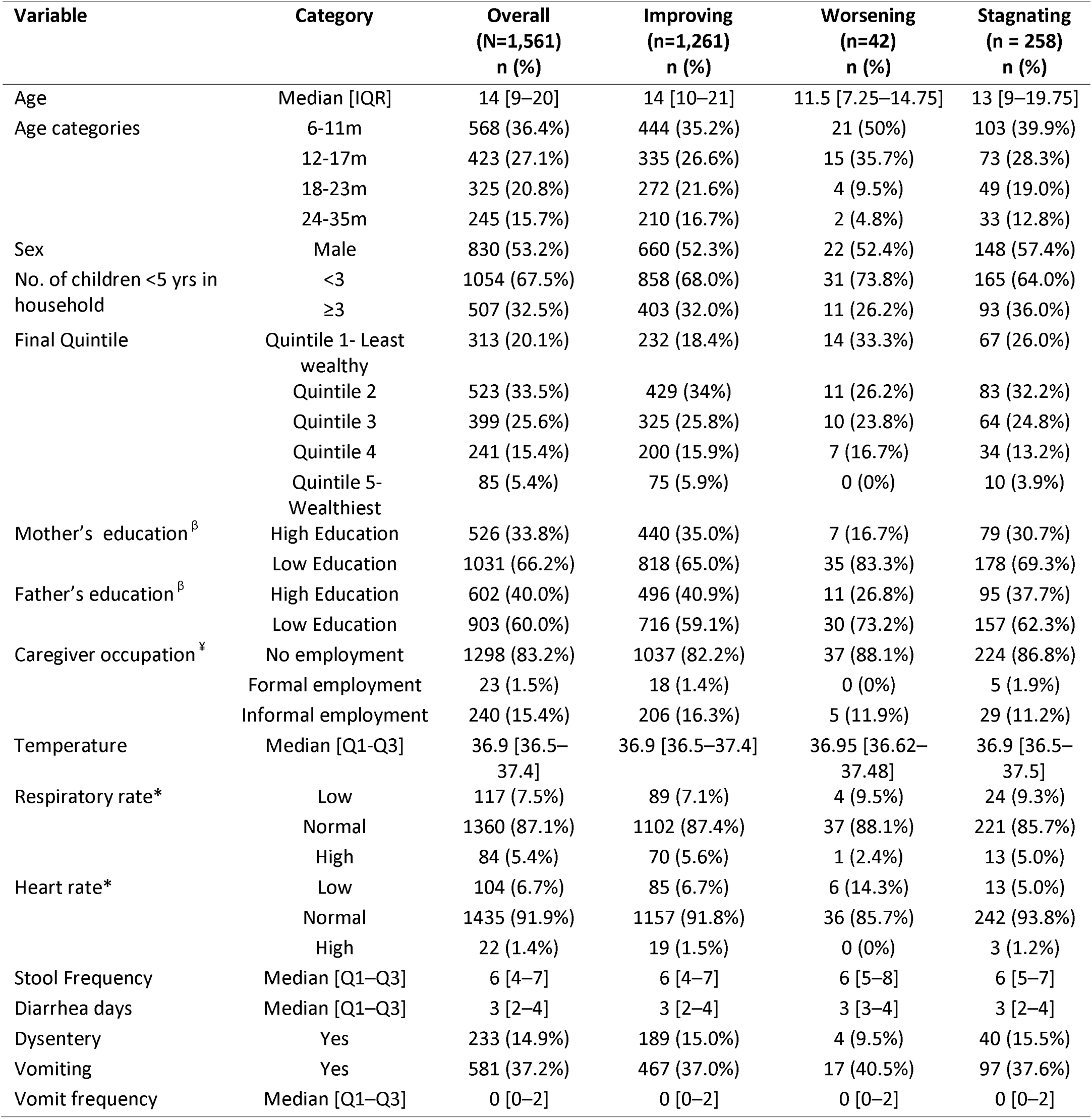
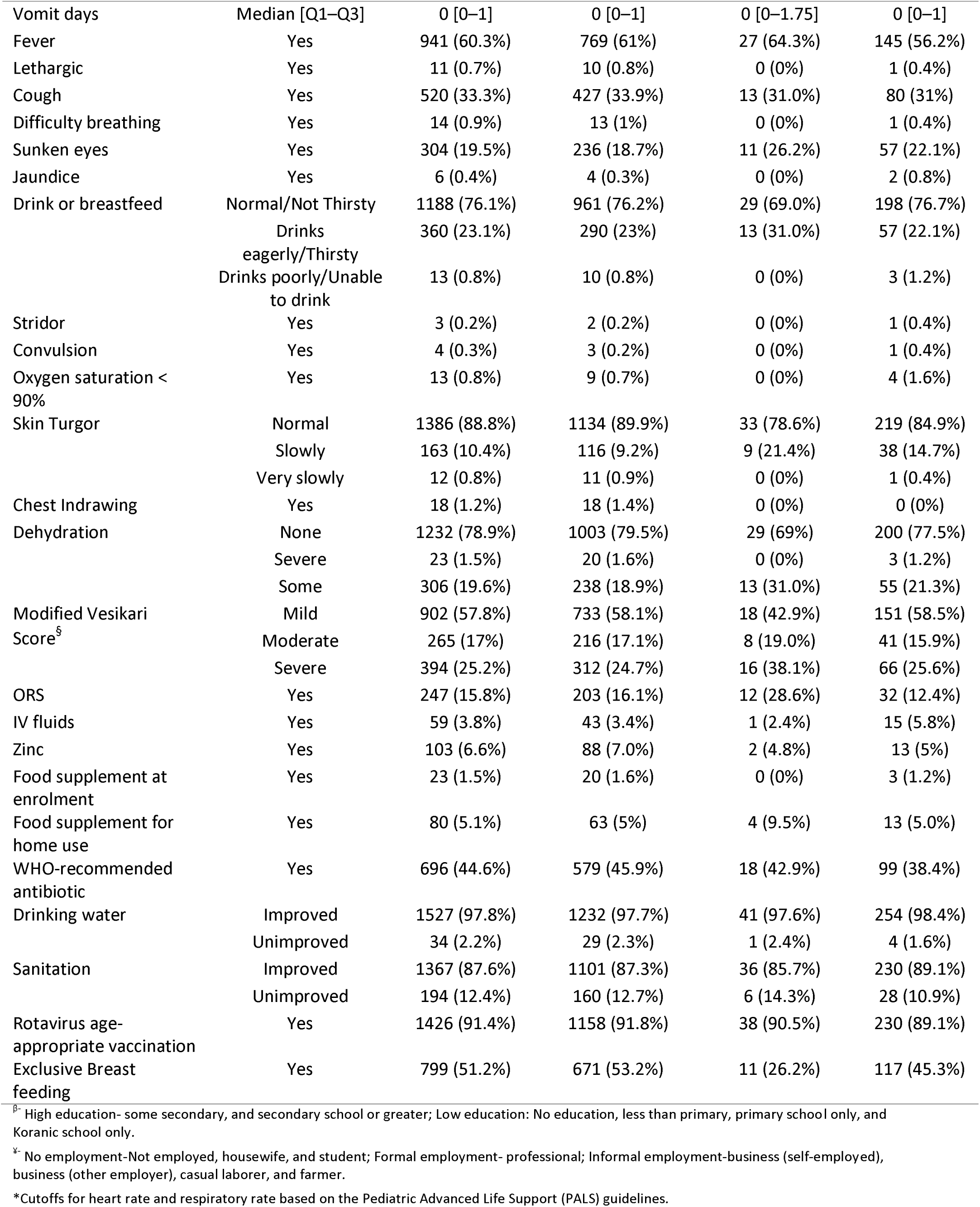
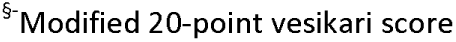
Baseline Characteristics of Wasted Children aged 6-35 months Stratified by Post-Diarrhea Nutritional Recovery Trajectories across EFGH sites, 2022-2024.

| Variable | Category | Overall<br>(N=1,561)<br>n (%) | Improving<br>(n=1,261)<br>n (%) | Worsening<br>(n=42)<br>n (%) | Stagnating<br>(n = 258)<br>n (%) |
| --- | --- | --- | --- | --- | --- |
| Age | Median [IQR] | 14 [9–20] | 14 [10–21] | 11.5 [7.25–14.75] | 13 [9–19.75] |
| Age categories | 6-11m | 568 (36.4%) | 444 (35.2%) | 21 (50%) | 103 (39.9%) |
|  | 12-17m | 423 (27.1%) | 335 (26.6%) | 15 (35.7%) | 73 (28.3%) |
|  | 18-23m | 325 (20.8%) | 272 (21.6%) | 4 (9.5%) | 49 (19.0%) |
|  | 24-35m | 245 (15.7%) | 210 (16.7%) | 2 (4.8%) | 33 (12.8%) |
| Sex | Male | 830 (53.2%) | 660 (52.3%) | 22 (52.4%) | 148 (57.4%) |
| No. of children <5 yrs in household | <3 | 1054 (67.5%) | 858 (68.0%) | 31 (73.8%) | 165 (64.0%) |
|  | ≥3 | 507 (32.5%) | 403 (32.0%) | 11 (26.2%) | 93 (36.0%) |
| Final Quintile | Quintile 1- Least wealthy | 313 (20.1%) | 232 (18.4%) | 14 (33.3%) | 67 (26.0%) |
|  | Quintile 2 | 523 (33.5%) | 429 (34%) | 11 (26.2%) | 83 (32.2%) |
|  | Quintile 3 | 399 (25.6%) | 325 (25.8%) | 10 (23.8%) | 64 (24.8%) |
|  | Quintile 4 | 241 (15.4%) | 200 (15.9%) | 7 (16.7%) | 34 (13.2%) |
|  | Quintile 5- Wealthiest | 85 (5.4%) | 75 (5.9%) | 0 (0%) | 10 (3.9%) |
| Mother's education<sup>β</sup> | High Education | 526 (33.8%) | 440 (35.0%) | 7 (16.7%) | 79 (30.7%) |
|  | Low Education | 1031 (66.2%) | 818 (65.0%) | 35 (83.3%) | 178 (69.3%) |
| Father's education<sup>β</sup> | High Education | 602 (40.0%) | 496 (40.9%) | 11 (26.8%) | 95 (37.7%) |
|  | Low Education | 903 (60.0%) | 716 (59.1%) | 30 (73.2%) | 157 (62.3%) |
| Caregiver occupation<sup>¥</sup> | No employment | 1298 (83.2%) | 1037 (82.2%) | 37 (88.1%) | 224 (86.8%) |
|  | Formal employment | 23 (1.5%) | 18 (1.4%) | 0 (0%) | 5 (1.9%) |
|  | Informal employment | 240 (15.4%) | 206 (16.3%) | 5 (11.9%) | 29 (11.2%) |
| Temperature | Median [Q1-Q3] | 36.9 [36.5–37.4] | 36.9 [36.5–37.4] | 36.95 [36.62–37.48] | 36.9 [36.5–37.5] |
| Respiratory rate* | Low | 117 (7.5%) | 89 (7.1%) | 4 (9.5%) | 24 (9.3%) |
|  | Normal | 1360 (87.1%) | 1102 (87.4%) | 37 (88.1%) | 221 (85.7%) |
|  | High | 84 (5.4%) | 70 (5.6%) | 1 (2.4%) | 13 (5.0%) |
| Heart rate* | Low | 104 (6.7%) | 85 (6.7%) | 6 (14.3%) | 13 (5.0%) |
|  | Normal | 1435 (91.9%) | 1157 (91.8%) | 36 (85.7%) | 242 (93.8%) |
|  | High | 22 (1.4%) | 19 (1.5%) | 0 (0%) | 3 (1.2%) |
| Stool Frequency | Median [Q1–Q3] | 6 [4–7] | 6 [4–7] | 6 [5–8] | 6 [5–7] |
| Diarrhea days | Median [Q1–Q3] | 3 [2–4] | 3 [2–4] | 3 [3–4] | 3 [2–4] |
| Dysentery | Yes | 233 (14.9%) | 189 (15.0%) | 4 (9.5%) | 40 (15.5%) |
| Vomiting | Yes | 581 (37.2%) | 467 (37.0%) | 17 (40.5%) | 97 (37.6%) |
| Vomit frequency | Median [Q1–Q3] | 0 [0–2] | 0 [0–2] | 0 [0–2] | 0 [0–2] |
| Vomit days | Median [Q1–Q3] | 0 [0–1] | 0 [0–1] | 0 [0–1.75] | 0 [0–1] |
| Fever | Yes | 941 (60.3%) | 769 (61%) | 27 (64.3%) | 145 (56.2%) |
| Lethargic | Yes | 11 (0.7%) | 10 (0.8%) | 0 (0%) | 1 (0.4%) |
| Cough | Yes | 520 (33.3%) | 427 (33.9%) | 13 (31.0%) | 80 (31%) |
| Difficulty breathing | Yes | 14 (0.9%) | 13 (1%) | 0 (0%) | 1 (0.4%) |
| Sunken eyes | Yes | 304 (19.5%) | 236 (18.7%) | 11 (26.2%) | 57 (22.1%) |
| Jaundice | Yes | 6 (0.4%) | 4 (0.3%) | 0 (0%) | 2 (0.8%) |
| Drink or breastfeed | Normal/Not Thirsty | 1188 (76.1%) | 961 (76.2%) | 29 (69.0%) | 198 (76.7%) |
|  | Drinks eagerly/Thirsty | 360 (23.1%) | 290 (23%) | 13 (31.0%) | 57 (22.1%) |
|  | Drinks poorly/Unable to drink | 13 (0.8%) | 10 (0.8%) | 0 (0%) | 3 (1.2%) |
| Stridor | Yes | 3 (0.2%) | 2 (0.2%) | 0 (0%) | 1 (0.4%) |
| Convulsion | Yes | 4 (0.3%) | 3 (0.2%) | 0 (0%) | 1 (0.4%) |
| Oxygen saturation < 90% | Yes | 13 (0.8%) | 9 (0.7%) | 0 (0%) | 4 (1.6%) |
| Skin Turgor | Normal | 1386 (88.8%) | 1134 (89.9%) | 33 (78.6%) | 219 (84.9%) |
|  | Slowly | 163 (10.4%) | 116 (9.2%) | 9 (21.4%) | 38 (14.7%) |
|  | Very slowly | 12 (0.8%) | 11 (0.9%) | 0 (0%) | 1 (0.4%) |
| Chest Indrawing | Yes | 18 (1.2%) | 18 (1.4%) | 0 (0%) | 0 (0%) |
| Dehydration | None | 1232 (78.9%) | 1003 (79.5%) | 29 (69%) | 200 (77.5%) |
|  | Severe | 23 (1.5%) | 20 (1.6%) | 0 (0%) | 3 (1.2%) |
|  | Some | 306 (19.6%) | 238 (18.9%) | 13 (31.0%) | 55 (21.3%) |
| Modified Vesikari Score<sup>5</sup> | Mild | 902 (57.8%) | 733 (58.1%) | 18 (42.9%) | 151 (58.5%) |
|  | Moderate | 265 (17%) | 216 (17.1%) | 8 (19.0%) | 41 (15.9%) |
|  | Severe | 394 (25.2%) | 312 (24.7%) | 16 (38.1%) | 66 (25.6%) |
| ORS | Yes | 247 (15.8%) | 203 (16.1%) | 12 (28.6%) | 32 (12.4%) |
| IV fluids | Yes | 59 (3.8%) | 43 (3.4%) | 1 (2.4%) | 15 (5.8%) |
| Zinc | Yes | 103 (6.6%) | 88 (7.0%) | 2 (4.8%) | 13 (5%) |
| Food supplement at enrolment | Yes | 23 (1.5%) | 20 (1.6%) | 0 (0%) | 3 (1.2%) |
| Food supplement for home use | Yes | 80 (5.1%) | 63 (5%) | 4 (9.5%) | 13 (5.0%) |
| WHO-recommended antibiotic | Yes | 696 (44.6%) | 579 (45.9%) | 18 (42.9%) | 99 (38.4%) |
| Drinking water | Improved | 1527 (97.8%) | 1232 (97.7%) | 41 (97.6%) | 254 (98.4%) |
|  | Unimproved | 34 (2.2%) | 29 (2.3%) | 1 (2.4%) | 4 (1.6%) |
| Sanitation | Improved | 1367 (87.6%) | 1101 (87.3%) | 36 (85.7%) | 230 (89.1%) |
|  | Unimproved | 194 (12.4%) | 160 (12.7%) | 6 (14.3%) | 28 (10.9%) |
| Rotavirus age-appropriate vaccination | Yes | 1426 (91.4%) | 1158 (91.8%) | 38 (90.5%) | 230 (89.1%) |
| Exclusive Breast feeding | Yes | 799 (51.2%) | 671 (53.2%) | 11 (26.2%) | 117 (45.3%) |

<sup>5</sup>- High education- some secondary, and secondary school or greater; Low education: No education, less than primary, primary school only, and Koranic school only.
<sup>6</sup>- No employment-Not employed, housewife, and student; Formal employment- professional; Informal employment-business (self-employed), business (other employer), casual laborer, and farmer.
\*Cutoffs for heart rate and respiratory rate based on the Pediatric Advanced Life Support (PALS) guidelines.
<sup>§</sup>-Modified 20-point vesikari score

**Table 3:**
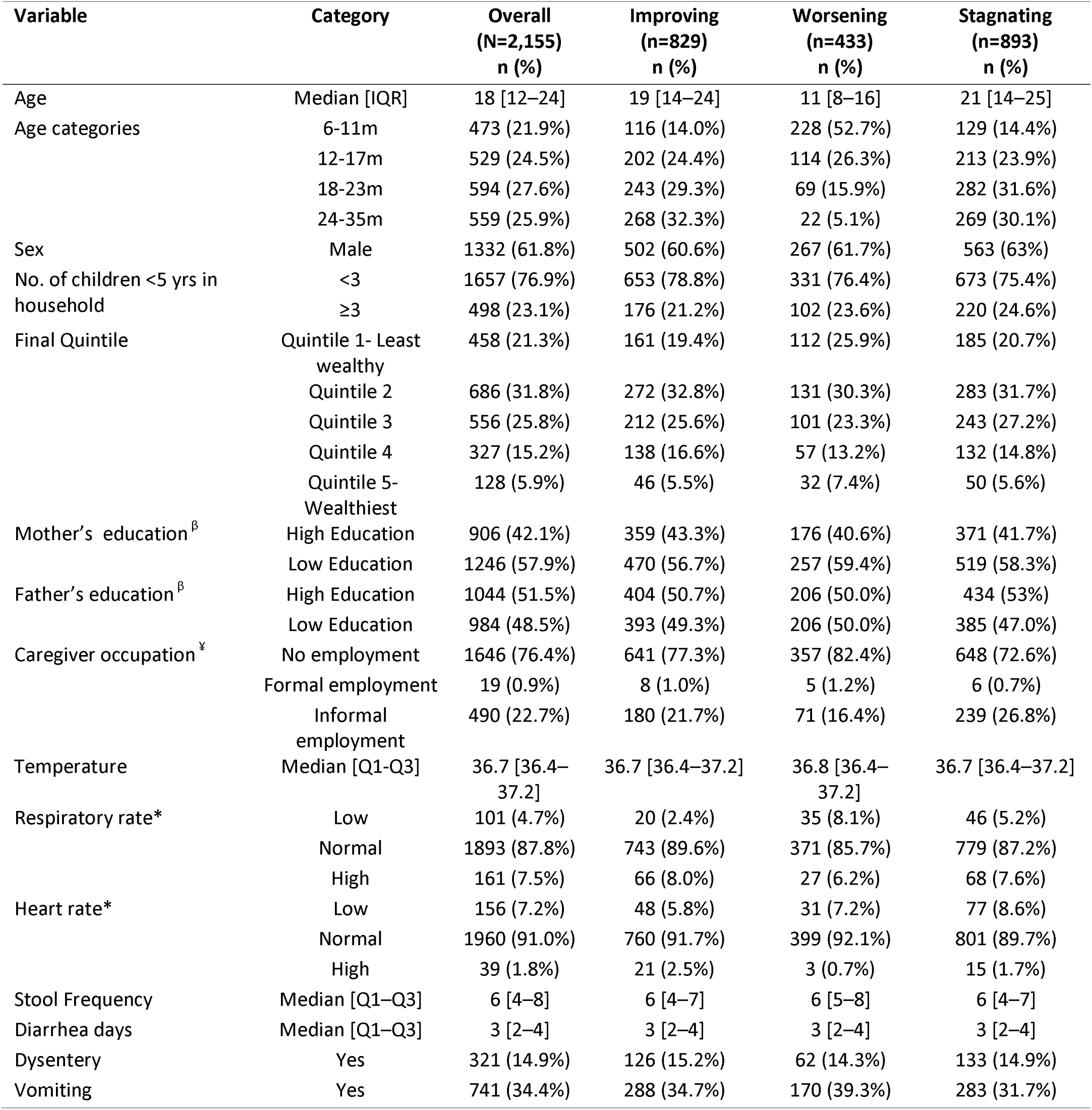
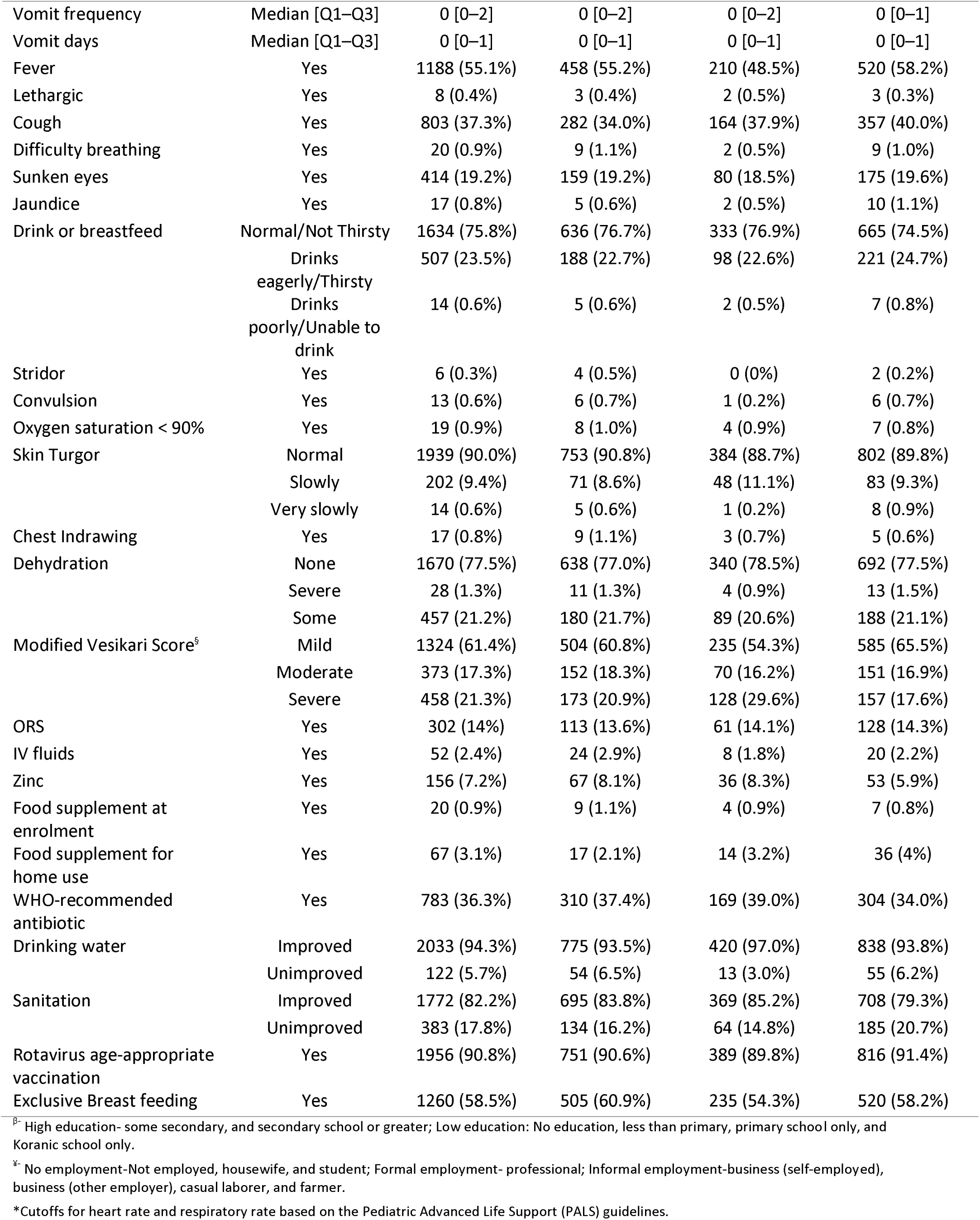
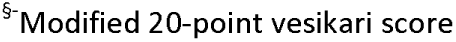
Baseline Characteristics of Stunted Children aged 6-35 months Stratified by Post-Diarrhea Nutritional Recovery Trajectories across EFGH sites, 2022-2024.

| Variable | Category | Overall<br>(N=2,155)<br>n (%) | Improving<br>(n=829)<br>n (%) | Worsening<br>(n=433)<br>n (%) | Stagnating<br>(n=893)<br>n (%) |
| --- | --- | --- | --- | --- | --- |
| Age | Median [IQR] | 18 [12–24] | 19 [14–24] | 11 [8–16] | 21 [14–25] |
| Age categories | 6-11m | 473 (21.9%) | 116 (14.0%) | 228 (52.7%) | 129 (14.4%) |
|  | 12-17m | 529 (24.5%) | 202 (24.4%) | 114 (26.3%) | 213 (23.9%) |
|  | 18-23m | 594 (27.6%) | 243 (29.3%) | 69 (15.9%) | 282 (31.6%) |
|  | 24-35m | 559 (25.9%) | 268 (32.3%) | 22 (5.1%) | 269 (30.1%) |
| Sex | Male | 1332 (61.8%) | 502 (60.6%) | 267 (61.7%) | 563 (63%) |
| No. of children <5 yrs in household | <3 | 1657 (76.9%) | 653 (78.8%) | 331 (76.4%) | 673 (75.4%) |
|  | ≥3 | 498 (23.1%) | 176 (21.2%) | 102 (23.6%) | 220 (24.6%) |
| Final Quintile | Quintile 1- Least wealthy | 458 (21.3%) | 161 (19.4%) | 112 (25.9%) | 185 (20.7%) |
|  | Quintile 2 | 686 (31.8%) | 272 (32.8%) | 131 (30.3%) | 283 (31.7%) |
|  | Quintile 3 | 556 (25.8%) | 212 (25.6%) | 101 (23.3%) | 243 (27.2%) |
|  | Quintile 4 | 327 (15.2%) | 138 (16.6%) | 57 (13.2%) | 132 (14.8%) |
|  | Quintile 5- Wealthiest | 128 (5.9%) | 46 (5.5%) | 32 (7.4%) | 50 (5.6%) |
| Mother's education<sup>§</sup> | High Education | 906 (42.1%) | 359 (43.3%) | 176 (40.6%) | 371 (41.7%) |
|  | Low Education | 1246 (57.9%) | 470 (56.7%) | 257 (59.4%) | 519 (58.3%) |
| Father's education<sup>§</sup> | High Education | 1044 (51.5%) | 404 (50.7%) | 206 (50.0%) | 434 (53%) |
|  | Low Education | 984 (48.5%) | 393 (49.3%) | 206 (50.0%) | 385 (47.0%) |
| Caregiver occupation<sup>§</sup> | No employment | 1646 (76.4%) | 641 (77.3%) | 357 (82.4%) | 648 (72.6%) |
|  | Formal employment | 19 (0.9%) | 8 (1.0%) | 5 (1.2%) | 6 (0.7%) |
|  | Informal employment | 490 (22.7%) | 180 (21.7%) | 71 (16.4%) | 239 (26.8%) |
| Temperature | Median [Q1-Q3] | 36.7 [36.4–37.2] | 36.7 [36.4–37.2] | 36.8 [36.4–37.2] | 36.7 [36.4–37.2] |
| Respiratory rate* | Low | 101 (4.7%) | 20 (2.4%) | 35 (8.1%) | 46 (5.2%) |
|  | Normal | 1893 (87.8%) | 743 (89.6%) | 371 (85.7%) | 779 (87.2%) |
|  | High | 161 (7.5%) | 66 (8.0%) | 27 (6.2%) | 68 (7.6%) |
| Heart rate* | Low | 156 (7.2%) | 48 (5.8%) | 31 (7.2%) | 77 (8.6%) |
|  | Normal | 1960 (91.0%) | 760 (91.7%) | 399 (92.1%) | 801 (89.7%) |
|  | High | 39 (1.8%) | 21 (2.5%) | 3 (0.7%) | 15 (1.7%) |
| Stool Frequency | Median [Q1-Q3] | 6 [4–8] | 6 [4–7] | 6 [5–8] | 6 [4–7] |
| Diarrhea days | Median [Q1-Q3] | 3 [2–4] | 3 [2–4] | 3 [2–4] | 3 [2–4] |
| Dysentery | Yes | 321 (14.9%) | 126 (15.2%) | 62 (14.3%) | 133 (14.9%) |
| Vomiting | Yes | 741 (34.4%) | 288 (34.7%) | 170 (39.3%) | 283 (31.7%) |
| Vomit frequency | Median [Q1–Q3] | 0 [0–2] | 0 [0–2] | 0 [0–2] | 0 [0–1] |
| Vomit days | Median [Q1–Q3] | 0 [0–1] | 0 [0–1] | 0 [0–1] | 0 [0–1] |
| Fever | Yes | 1188 (55.1%) | 458 (55.2%) | 210 (48.5%) | 520 (58.2%) |
| Lethargic | Yes | 8 (0.4%) | 3 (0.4%) | 2 (0.5%) | 3 (0.3%) |
| Cough | Yes | 803 (37.3%) | 282 (34.0%) | 164 (37.9%) | 357 (40.0%) |
| Difficulty breathing | Yes | 20 (0.9%) | 9 (1.1%) | 2 (0.5%) | 9 (1.0%) |
| Sunken eyes | Yes | 414 (19.2%) | 159 (19.2%) | 80 (18.5%) | 175 (19.6%) |
| Jaundice | Yes | 17 (0.8%) | 5 (0.6%) | 2 (0.5%) | 10 (1.1%) |
| Drink or breastfeed | Normal/Not Thirsty | 1634 (75.8%) | 636 (76.7%) | 333 (76.9%) | 665 (74.5%) |
|  | Drinks eagerly/Thirsty | 507 (23.5%) | 188 (22.7%) | 98 (22.6%) | 221 (24.7%) |
|  | Drinks poorly/Unable to drink | 14 (0.6%) | 5 (0.6%) | 2 (0.5%) | 7 (0.8%) |
| Stridor | Yes | 6 (0.3%) | 4 (0.5%) | 0 (0%) | 2 (0.2%) |
| Convulsion | Yes | 13 (0.6%) | 6 (0.7%) | 1 (0.2%) | 6 (0.7%) |
| Oxygen saturation < 90% | Yes | 19 (0.9%) | 8 (1.0%) | 4 (0.9%) | 7 (0.8%) |
| Skin Turgor | Normal | 1939 (90.0%) | 753 (90.8%) | 384 (88.7%) | 802 (89.8%) |
|  | Slowly | 202 (9.4%) | 71 (8.6%) | 48 (11.1%) | 83 (9.3%) |
|  | Very slowly | 14 (0.6%) | 5 (0.6%) | 1 (0.2%) | 8 (0.9%) |
| Chest Indrawing | Yes | 17 (0.8%) | 9 (1.1%) | 3 (0.7%) | 5 (0.6%) |
| Dehydration | None | 1670 (77.5%) | 638 (77.0%) | 340 (78.5%) | 692 (77.5%) |
|  | Severe | 28 (1.3%) | 11 (1.3%) | 4 (0.9%) | 13 (1.5%) |
|  | Some | 457 (21.2%) | 180 (21.7%) | 89 (20.6%) | 188 (21.1%) |
| Modified Vesikari Score<sup>§</sup> | Mild | 1324 (61.4%) | 504 (60.8%) | 235 (54.3%) | 585 (65.5%) |
|  | Moderate | 373 (17.3%) | 152 (18.3%) | 70 (16.2%) | 151 (16.9%) |
|  | Severe | 458 (21.3%) | 173 (20.9%) | 128 (29.6%) | 157 (17.6%) |
| ORS | Yes | 302 (14%) | 113 (13.6%) | 61 (14.1%) | 128 (14.3%) |
| IV fluids | Yes | 52 (2.4%) | 24 (2.9%) | 8 (1.8%) | 20 (2.2%) |
| Zinc | Yes | 156 (7.2%) | 67 (8.1%) | 36 (8.3%) | 53 (5.9%) |
| Food supplement at enrolment | Yes | 20 (0.9%) | 9 (1.1%) | 4 (0.9%) | 7 (0.8%) |
| Food supplement for home use | Yes | 67 (3.1%) | 17 (2.1%) | 14 (3.2%) | 36 (4%) |
| WHO-recommended antibiotic | Yes | 783 (36.3%) | 310 (37.4%) | 169 (39.0%) | 304 (34.0%) |
| Drinking water | Improved | 2033 (94.3%) | 775 (93.5%) | 420 (97.0%) | 838 (93.8%) |
|  | Unimproved | 122 (5.7%) | 54 (6.5%) | 13 (3.0%) | 55 (6.2%) |
| Sanitation | Improved | 1772 (82.2%) | 695 (83.8%) | 369 (85.2%) | 708 (79.3%) |
|  | Unimproved | 383 (17.8%) | 134 (16.2%) | 64 (14.8%) | 185 (20.7%) |
| Rotavirus age-appropriate vaccination | Yes | 1956 (90.8%) | 751 (90.6%) | 389 (89.8%) | 816 (91.4%) |
| Exclusive Breast feeding | Yes | 1260 (58.5%) | 505 (60.9%) | 235 (54.3%) | 520 (58.2%) |

<sup>§</sup> High education- some secondary, and secondary school or greater; Low education: No education, less than primary, primary school only, and Koranic school only.
<sup>¶</sup> No employment-Not employed, housewife, and student; Formal employment- professional; Informal employment-business (self-employed), business (other employer), casual laborer, and farmer.
\*Cutoffs for heart rate and respiratory rate based on the Pediatric Advanced Life Support (PALS) guidelines.
<sup>§</sup>-Modified 20-point vesikari score

**Table 4:**
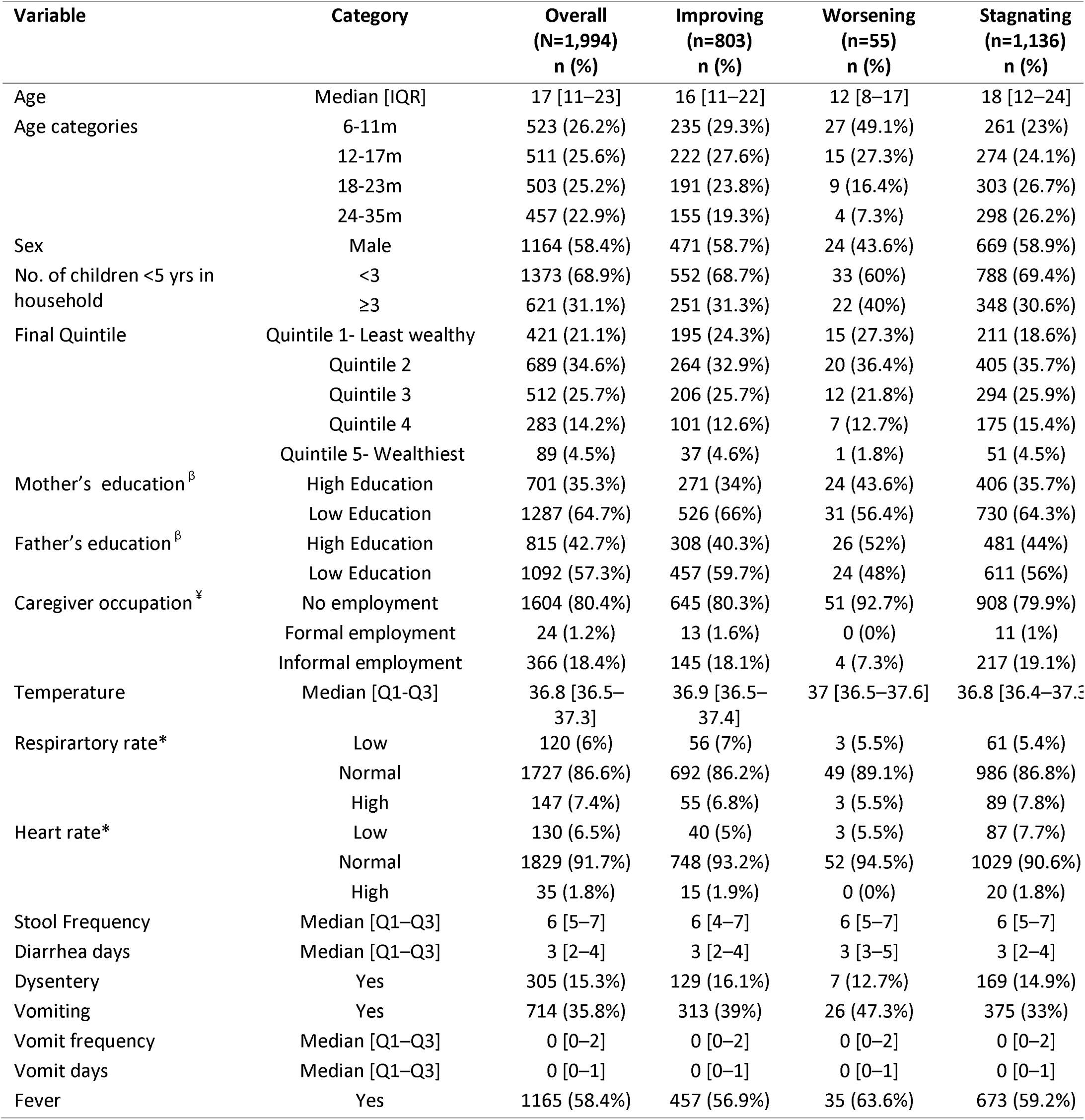
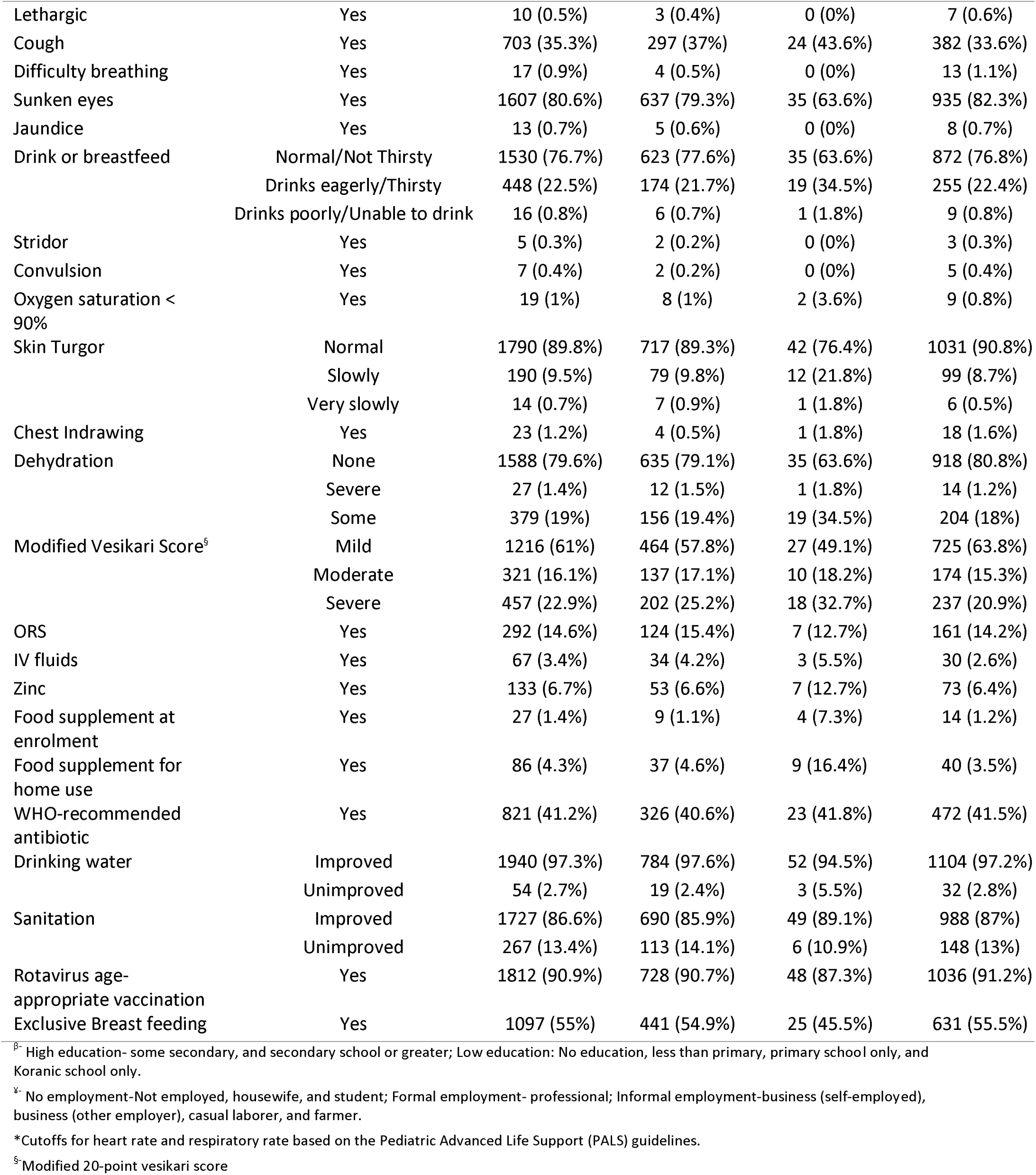
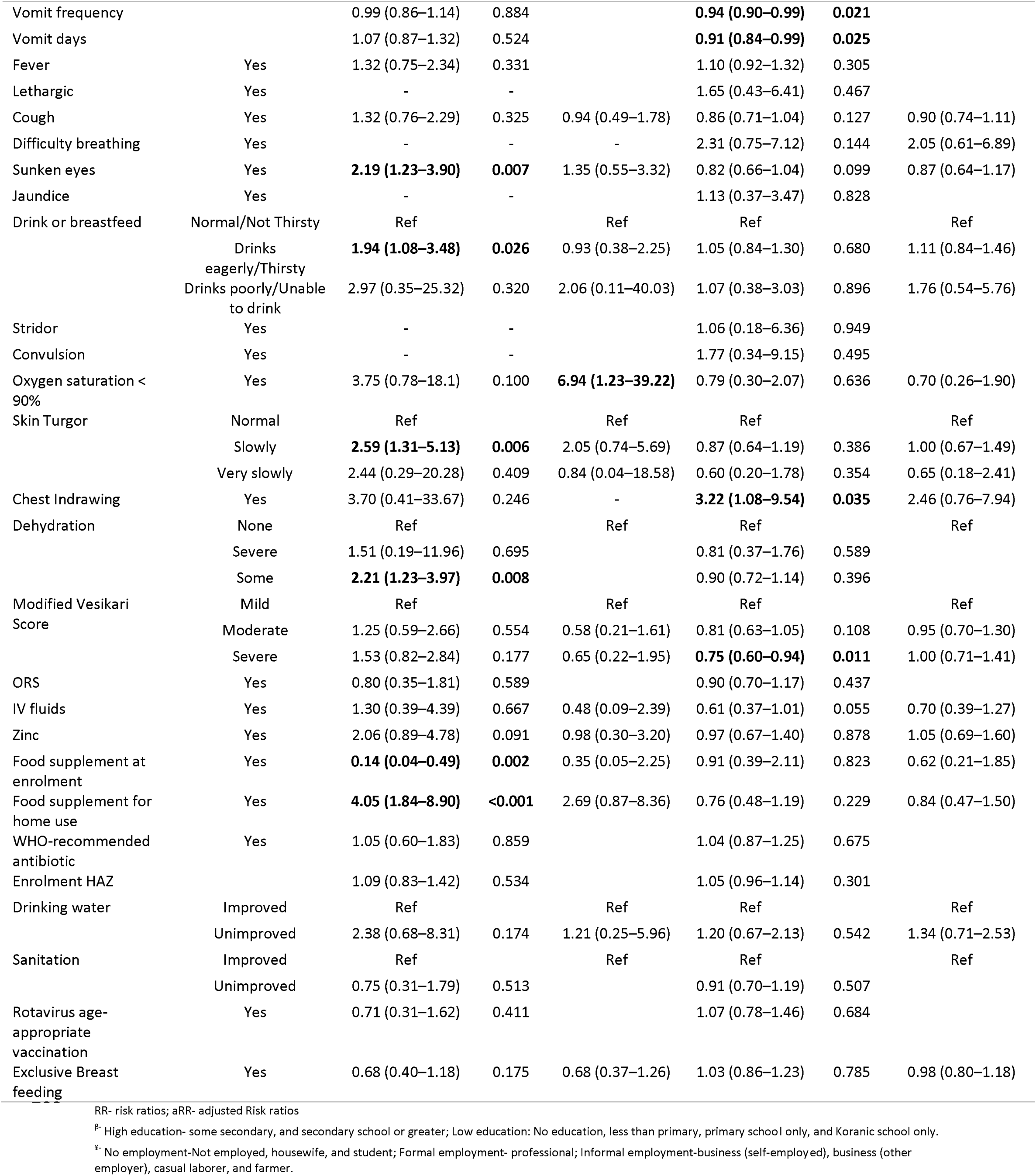
Baseline Characteristics of Underweight Children aged 6-35 months Stratified by Post-Diarrhea Nutritional Recovery Trajectories across EFGH sites, 2022-2024.

Despite diverging outcomes, baseline anthropometric scores were comparable across the three eventual trajectories within each cohort (Figure 2). At enrollment, median WHZ for the wasting cohort ranged from -2.14 to -2.37; median HAZ for the stunted cohort ranged from -2.56 to -2.67; and median WAZ for the underweight cohort ranged from -2.54 to -2.74. No significant differences in baseline anthropometric severity were observed between children who later improved, stagnated, or worsened (Figure 2).

**Figure 2.**
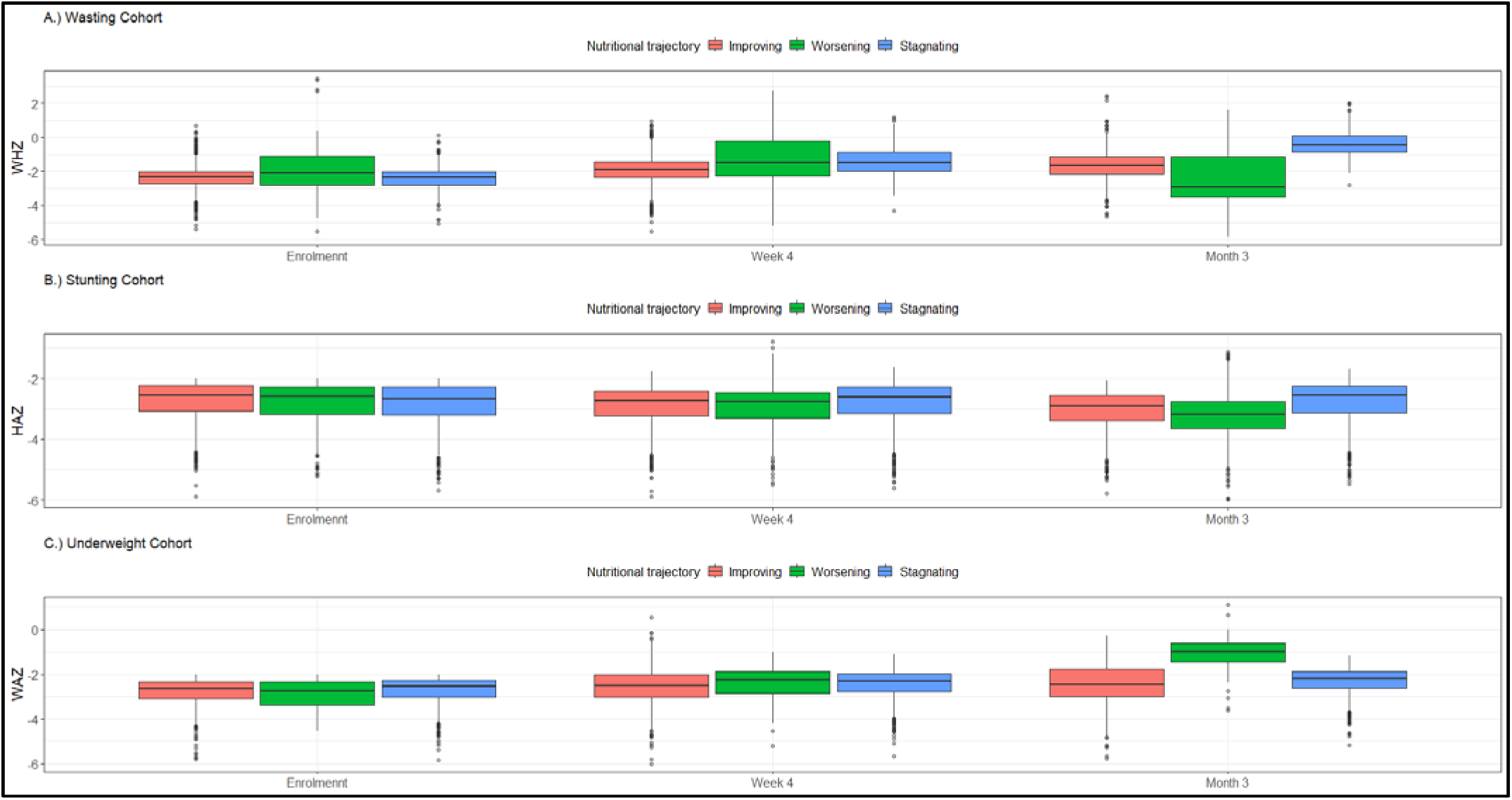
Baseline and Follow-up Anthropometric Status in the Wasting, Stunting and Underweight Cohorts Stratified by Nutritional trajectories

### Factors Associated with Nutritional Trajectories Wasting Cohort

In multinomial modified Poisson regression analyses with improving nutritional trajectory as the reference category, several sociodemographic, and clinical factors were differentially associated with worsening and stagnating trajectories among wasted children following a diarrheal illness. Among children in the worsening trajectory, exclusive breastfeeding and age were the only significant protective factors; children who were exclusively breastfed had 63% lower risk of experiencing a worsening trajectory compared to those who were not (aRR: 0.37, 95% CI: 0.18–0.77) while those aged 18-23 months had a 72% lower risk of stagnation (aRR: 0.28, 95% CI: 0.09–0.86) compared to those age 6-11 months.Clinical markers of severity at enrollment were strong predictors of stagnation; specifically, each additional day of diarrhea duration increased the risk of stagnation by 11% (aRR: 1.11, 95% CI: 1.01–1.23), and children with slow skin turgor (a sign of dehydration) were nearly twice as likely to stagnate compared to those with normal turgor (aRR: 1.96, 95% CI: 1.18–3.27) (Figure 3, Table S2).

**Figure 3.**
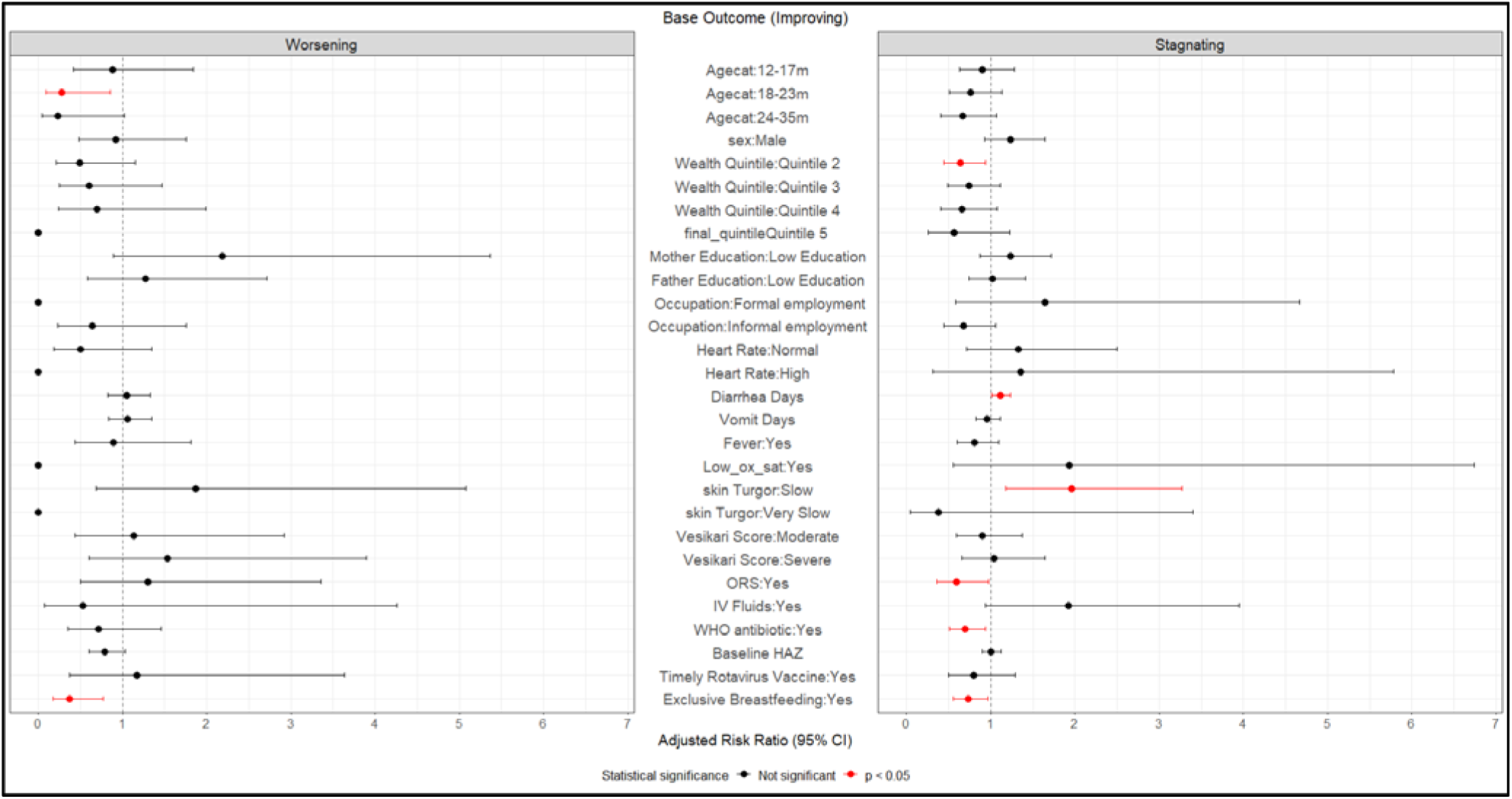
Factors Associated with Post-diarrheal Worsening and Stagnating Nutritional Trajectories Among Children Wasted at Baseline.

Conversely, certain interventions and household characteristics were protective against stagnation. Children from households in the second wealth quintile had a 36% lower risk of stagnation 0.64 (0.44–0.94). Regarding clinical management, the use of Oral Rehydration Salts (ORS) (aRR: 0.59, 95% CI: 0.36–0.97) and the administration of WHO-recommended antibiotics (aRR: 0.69, 95% CI: 0.51–0.94) were both significantly associated with a reduced risk of stagnation. Furthermore, exclusive breastfeeding remained protective for this group, reducing the risk of stagnation by 28% (aRR: 0.73, 95% CI: 0.55–0.96) (Figure 3, Table S2).

### Stunting Cohort

In the stunting cohort, multinomial regression analysis revealed that the drivers of nutritional trajectories were distinctly different from those observed in the wasting cohort, (Figure 4, Table S3). The risk of a worsening nutritional trajectory was most strongly associated with younger age. Compared to infants (6–11 months), children in older age categories were significantly less likely to experience worsening stunting, with the risk decreasing drastically for those aged 12–17 months (aRR: 0.29, 95% CI: 0.21–0.41), 18–23 months (aRR: 0.14, 95% CI: 0.10–0.21), and 24–35 months (aRR: 0.04, 95% CI: 0.02–0.06). Residing in wealthier quintiles were both protective against worsening (Quintile 3: aRR: 0.68, 95% CI: 0.46–0.99; Quintile 4: aRR: 0.50, 95% CI: 0.32–0.79). Unexpectedly, children with fever (aRR: 0.66, 95% CI: 0.50–0.87), low respiratory rate (aRR: 0.50, 95% CI: 0.27–0.94) , using unimproved drinking water (aRR: 0.39, 95% CI: 0.20–0.77) also showed a lower risk of worsening relative to the improving group.

**Figure 4.**
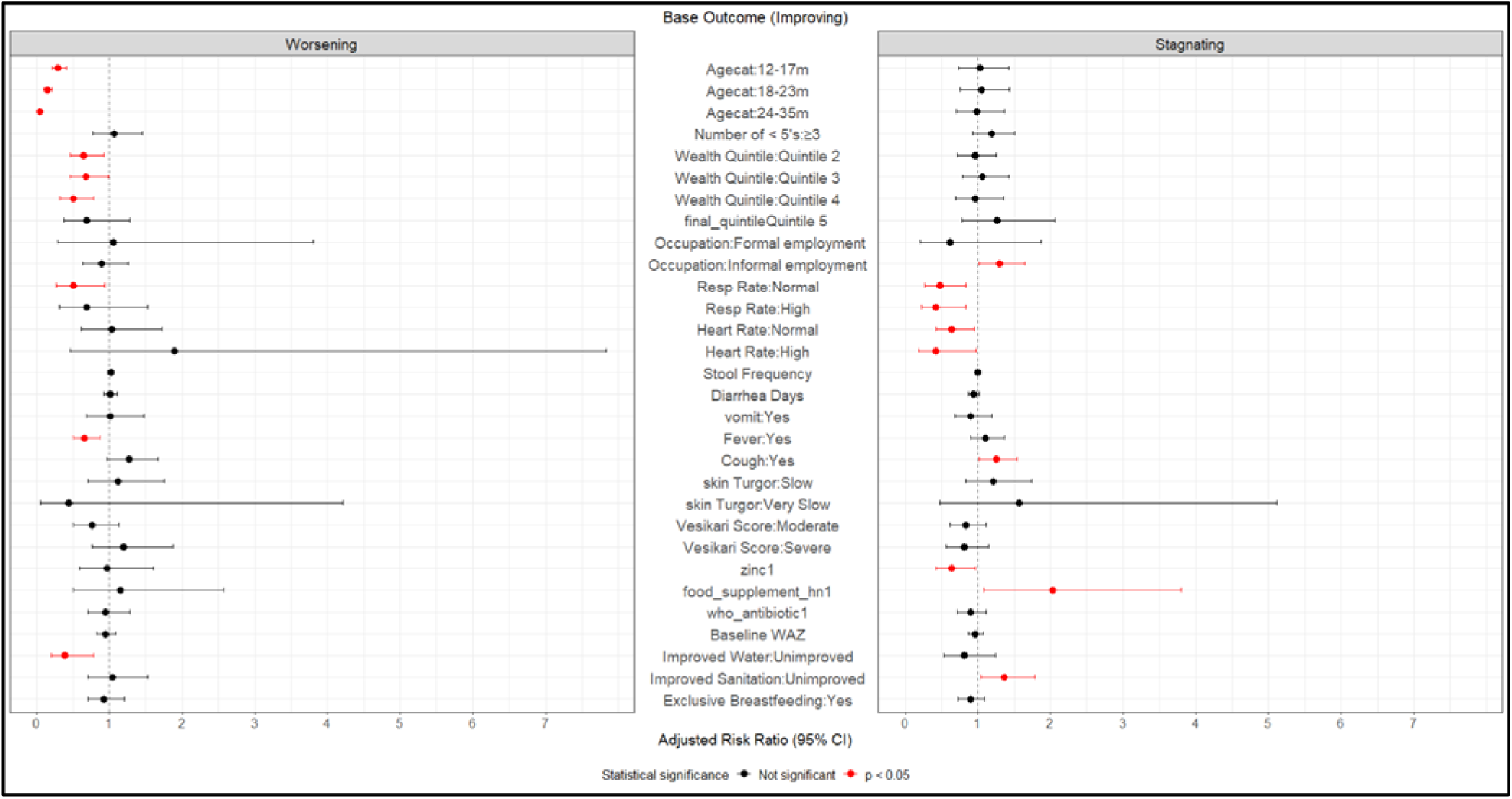
Factors Associated with Post-diarrheal Worsening and Stagnating Nutritional Trajectories Among Children Stunted at Baseline.

Children from households with unimproved sanitation were 36% more likely to stagnate (aRR: 1.36, 95% CI: 1.04–1.78). Caregiver employment also played a role, with informal employment associated with an increased risk of stagnation (aRR: 1.30, 95% CI: 1.02–1.63). Clinically, the presence of a cough at enrollment increased the risk of stagnation (aRR: 1.25, 95% CI: 1.02–1.54). Children receiving food supplements for home use were twice as likely to remain in the stagnating trajectory (aRR: 2.03, 95% CI: 1.08–3.980). Conversely, Abnormal vital signs, including low/high respiratory rates and low/high heart rates, were associated with a reduced risk of stagnation relative to the improving group. In terms of treatment, zinc supplementation was protective against stagnation (aRR: 0.64, 95% CI: 0.43–0.96) (Figure 4, Table S3).

### Underweight Cohort

For the final cohort of children who were underweight at enrollment, multinomial regression analysis revealed that the factors influencing nutritional decline (worsening) and lack of progress (stagnation) differed significantly, with clinical severity driving the former and age and socioeconomic status driving the latter (Figure 5, Table S4). Similar to the stunting cohort, infants (6–11 months) were at the highest risk; children in older age groups were significantly less likely to worsen, with the lowest risk among those aged 24–35 months (aRR: 0.23, 95% CI: 0.07–0.74). Sex was also a significant factor, as male children had a 54% lower risk of worsening compared to females (aRR: 0.46, 95% CI: 0.25–0.85). Children presenting with low oxygen saturation (<90%) had nearly a seven-fold increase in the risk of worsening (aRR: 6.94, 95% CI: 1.23–39.22). Furthermore, each additional day of diarrhea at enrollment was associated with a 24% increase in the risk of a worsening trajectory (aRR: 1.24, 95% CI: 1.02–1.52). Household dynamics also played a role, as the presence of ≥ 3 children < 5 years in the home nearly doubled the risk of worsening (aRR: 1.92, 95% CI: 1.01–3.64).

**Figure 5.**
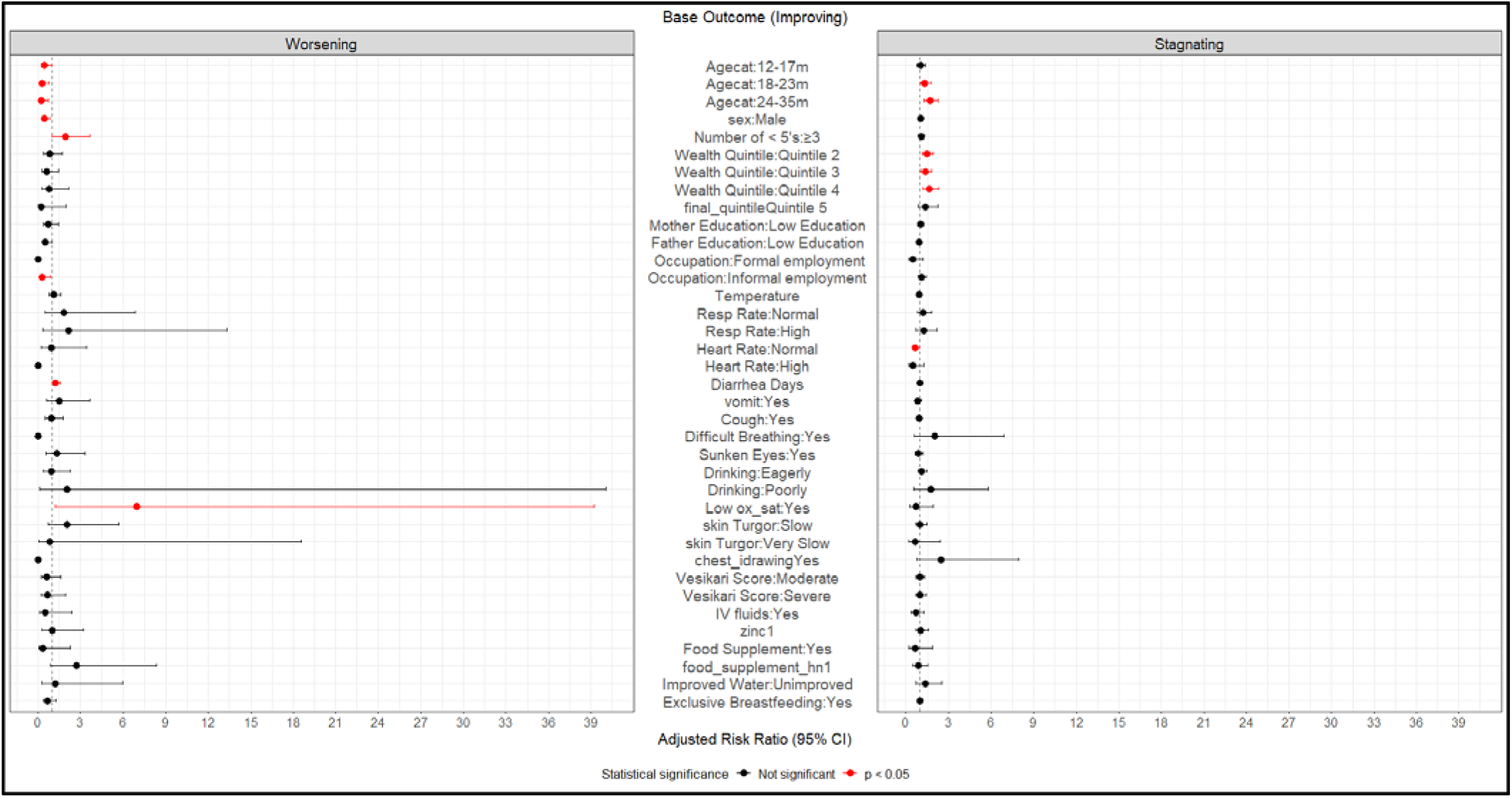
Factors Associated with Post-diarrheal Worsening and Stagnating Nutritional Trajectories Among Children Underweight at Baseline.

In contrast to the worsening trajectory, the risk of stagnation increased with age. Children aged 24–35 months were 70% more likely to stagnate rather than improve compared to infants (aRR: 1.70, 95% CI: 1.27–2.27). Socioeconomic status also showed an unexpected association: children from households in the second (aRR: 1.49), third (aRR: 1.37), and fourth (aRR: 1.64) wealth quintiles were all significantly more likely to experience stagnation compared to those in the lowest (least wealthy) quintile. Children presenting with a low heart rate were 37% less likely to stagnate (aRR: 0.63, 95% CI: 0.41–0.96) (Figure 5, Table S4).

## Discussion

Leveraging data from the EFGH Shigella surveillance study, we identified divergent post-diarrheal nutritional trajectories among children aged 6-35 months in a context of socioeconomic disadvantage. While wasting demonstrated substantial recovery potential, with four-fifths of affected children recovering, stunting and underweight proved far more entrenched, with fewer than two-fifths recovering and growth stagnation emerging as the predominant outcome for over half of these children. Moreover, the severity of baseline anthropometric deficits failed to predict future trajectories, revealing a paradox where identical initial presentations masked different biological and developmental pathways. Instead, we found that recovery was shaped by a confluence of factors operating at multiple levels: acute clinical markers of illness severity (prolonged diarrhea, dehydration, hypoxemia) increased the risk of nutritional failure. Age also emerged as a key determinant, with infants facing acute worsening while older toddlers confronted permanent stagnation. At the same time, modifiable protective factors including exclusive breastfeeding, ORS, appropriate antibiotics, and zinc supplementation, significantly improved outcomes, while unimproved sanitation persistently undermined them. These findings highlighting the imperative for integrated strategies that combine infection control, nutritional rehabilitation, and broader water, sanitation, and hygiene (WASH) and social protection interventions tailored to a child’s developmental stage.

More specifically, we observed divergent recovery patterns across the three nutritional cohorts. Wasting showed the greatest potential for recovery; four-fifths of wasted children recovered, compared to just two-fifths of those who were stunted or underweight. This recovery occurred against a backdrop of substantial socioeconomic disadvantage, characterized by high caregiver unemployment and low household wealth. This may partly reflect the influence of ongoing government initiatives in Kenya, such as community-based preventive health programs [24] and free healthcare services for children under five years [25], which may help mitigate the effects of household poverty by improving access to care. However, sustained recovery likely requires integrated strategies that combine clinical management and nutritional rehabilitation with broader social protection and livelihood support for vulnerable households. Growth stagnation emerged as a predominant trajectory among children who were stunted or underweight at enrolment, affecting between 41.4%-57.0% of the children. Worsening nutritional status was relatively uncommon, occurring in 2.7% of wasted and underweight children and in 20.1% of stunted children. These observations are broadly consistent with findings from previous studies that reported recovery rates of 27.0%-45.0% for stunting [26–28], 71.2% (95% CI: 68.5–73.8) for wasting [29] and 34% for underweight [30]. The similarity between these estimates and those observed in our cohort suggests that that diarrhea may not uniquely impair the biological mechanisms of nutritional recovery compared to other causes of undernutrition and that recovery following diarrheal illness may not differ substantially from recovery patterns reported for other causes. These results highlight the need for integrated post-diarrheal care strategies that combine infection management with nutritional rehabilitation and broader interventions targeting the determinants of chronic undernutrition.

Additionally, we observed a baseline paradox: the severity of anthropometric deficits at enrolment did not distinguish subsequent growth trajectories, as median WHZ, HAZ, and WAZ values were comparable among children who later improved, stagnated, or worsened. This finding aligns with observations by Dah et al., who reported poorer anthropometric and clinical outcomes among hospitalized children in Burkina Faso only when both WHZ and MUAC were used for case identification, compared with a single anthropometric criterion [31]. Together, these findings suggest that the depth of the initial growth deficit alone may be insufficient to predict post-diarrheal recovery as identical baseline scores may mask entirely different biological and developmental realities. Instead, recovery trajectories likely reflect a complex interplay of biological, environmental, and caregiving factors unfolding during and after the illness episode. These results underscore the limitations of relying on a single baseline measurement for risk stratification and highlight the need for more dynamic approaches to post-diarrheal nutritional management. Importantly, they also caution against prognostic bias in clinical care, reinforcing the need for consistent monitoring and treatment intensity for all undernourished children regardless of the severity of anthropometric deficits at presentation.

We also observed acute clinical markers at enrollment to be strong predictors of long-term nutritional failure. More specifically, prolonged diarrhea duration and dehydration (slow skin turgor) nearly doubled the risk of stagnation in wasted children. Prolonged diarrheal illness and dehydration can exacerbate nutrient losses, impair intestinal absorption, and increase metabolic demands during a period when children are already nutritionally vulnerable [32,33]. Similarly, low oxygen saturation (<90%) was associated with a seven-fold increase in the risk of nutritional decline, while each additional day of diarrhea increased the risk of worsening by 24% among underweight children. Together, these factors may limit the capacity for catch-up growth following illness and highlight that the severity and duration of concurrent illnesses play a critical role in shaping post-diarrheal nutritional trajectories. This underscores the importance of early detection and integrated management of both diarrheal disease and other acute infections.

Moreover, age emerged as a primary determinant of growth failure, with infancy identified as a critical window of vulnerability. Specifically, infants aged 6–11 months faced the highest risk of worsening trajectory in both stunting and underweight children. This is likely reflection of the convergence of several risk factors during this developmental stage, including rapid growth demands, the transition to complementary feeding, waning maternal antibodies and increased exposure to enteric pathogens [34]. Conversely, while the risk of acute worsening declined for older toddlers (24–35 months), this period was marked by an elevated probability of permanent stagnation, particularly among stunted or underweight children. This is possibly due to the body adapting physiologically to chronic undernutrition by slowing linear growth to conserve energy for survival [35,36]. While they are less likely to succumb acutely (worsen rapidly), their growth plates have likely undergone epigenetic changes that make catch-up growth metabolically difficult, leading to stagnation. Our findings reinforce the dogma that the first 1000 days are the critical window for preventing stunting. These findings highlight the need for age-stratified intervention strategies as recommended by UNICEF [37]. For infants, interventions should be preventive and proactive to rapidly identify and mitigate early growth declines. Priorities may include promoting safe weaning practices, improving WASH conditions, and high-quality complementary. In contrast, interventions for toddlers should emphasize sustained therapeutic support to address entrenched growth deficits. This may include the provision of more nutrient-dense therapeutic foods, integration of developmental stimulation and play-based interventions, and the management of underlying or subclinical infections such as routine deworming, to help break cycles of persistent growth stagnation.

We also identified several modifiable protective factors and interventions associated with improved post-diarrheal nutritional trajectories. First, exclusive breastfeeding remained the most robust protective factor across the nutritional cohorts consistent with existing literature [38]. The protective effect of exclusive breastfeeding is likely mediated through multiple mechanisms, including improved nutrient intake, enhanced immune protection through bioactive components in breast milk (IgA antibodies, lactoferrin, oligosaccharides), a healthy gut microbiome, and reduced exposure to contaminated foods and water [38]. In settings with high burdens of diarrheal disease and undernutrition, promoting exclusive breastfeeding remains one of the most cost-effective interventions for preventing growth faltering and supporting recovery. Consequently, interventions should prioritize maternal support systems (lactation counseling, workplace accommodations, protection from commercial pressures and adequate maternal nutritional support) to sustain exclusive breastfeeding. Second, the use of ORS and WHO-recommended antibiotics significantly reduced the risk of stagnation in wasted children, while zinc supplementation was protective against stagnation in the stunting cohort. This finding likely reflects the role of effective clinical management of diarrheal illness in minimizing dehydration, intestinal damage, and prolonged infection, factors that can exacerbate nutrient losses and impair recovery in already vulnerable children. Zinc may confer additional protection through its role in supporting intestinal mucosal repair, immune function, and improved nutrient absorption [39].

Finally, unimproved sanitation increased the risk of persistent stunting by 34%, highlighting the role of environmental conditions in shaping post-diarrheal recovery. Poor sanitation likely sustains repeated exposure to enteric pathogens, contributing to chronic intestinal inflammation, impaired nutrient absorption, and recurrent infections that limit the potential for catch-up growth [40,41]. These findings emphasize that clinical and nutritional interventions alone may be insufficient in settings with inadequate environmental health conditions. Strengthening WASH infrastructure, alongside clinical management and nutritional support, will be essential to break the cycle of infection and growth faltering and to support sustained recovery among vulnerable children.

Our study is not without limitations, which should be considered when interpreting the findings. First, the EFGH Shigella surveillance study was not originally designed or powered to specifically characterize post-diarrheal nutritional recovery. As a result, although the dataset provides rich longitudinal information, some determinants of growth, such as detailed dietary intake or household-level caregiving dynamics, may not have been fully captured. Second, anthropometric follow-up extended to three months after enrolment, which may be insufficient to fully capture longer-term recovery or chronic growth trajectories, particularly for linear growth deficits such as stunting. Third, LCMM assumes relative homogeneity within trajectory classes and may misclassify some individuals, especially within smaller groups. In addition, collapsing latent classes into broader clinical categories to enhance interpretability may obscure some finer distinctions in growth patterns. Nevertheless, the large multi-country prospective design, standardized anthropometric procedures, rigorous quality control procedures, and repeated follow-up measurements enhance confidence in the validity and robustness of the observed growth trajectory patterns.

## Conclusion

This study challenges the assumption that resolution of acute diarrhea equates to nutritional recovery, demonstrating instead that for stunted and underweight children, the dominant post-illness trajectory is growth stagnation. The discovery of a baseline paradox, where initial anthropometry fails to predict future decline, exposes a critical blind spot in current triage protocols, necessitating a shift from static screening to dynamic, longitudinal growth monitoring. Furthermore, the distinct vulnerabilities observed between infancy (acute deterioration) and toddlerhood (entrenched stagnation) buttresses the need to abandon one-size-fits-all approaches in favor of age-stratified precision public health strategies. Ultimately, biological recovery cannot be isolated from environmental context; the protective power of breastfeeding and clinical management was evident, yet often constrained by socioeconomic fragility and poor sanitation. Therefore, policy must pivot toward an integrated continuum of care that dissolves the silos between acute disease management, nutritional rehabilitation, and social protection. Only by coupling clinical excellence with structural support can health systems ensure that children not only survive diarrheal episodes but successfully regain their developmental potential.

## Acknowledgements

We extend our sincere gratitude to the study participants and their families across all sites for their willingness to participate in the EFGH study. We also thank the EFGH study staff for their dedication and hard work, which made this research possible. We are further grateful to the physicians, administrators, and Ministry of Health officials at recruitment sites in each participating country for providing facilities, operational support, and the institutional permissions that enabled successful study implementation.

The findings and conclusions presented in this report are those of the authors and do not necessarily represent the official positions of the Kenya Medical Research Institute or the partnering institutions.

## Data Availability

The EFGH data is publicly available through the Vivli data-sharing platform (https://search.vivli.org/doiLanding/studies/PR00011860/isLanding).

---

**Table S1:** Optimal Nutritional Trajectory Class Selection for Wasting, Stunting and Underweight using Latent Class Mixed-Effects Modeling.

| Nutritional Outcome | No. of Classes | loglik | AIC | BIC | Entropy |
| --- | --- | --- | --- | --- | --- |
| Wasting<br>(WHZ< -2 or MUAC<br><12.5) | 1 | -3124.41 | 6280.82 | 6366.448 | - |
|  | 2 | -3030.48 | 6100.967 | 6208.003 | 0.187908 |
|  | 3 | -2950.43 | 5948.859 | 6077.303 | 0.183977 |
|  | **4** | **-2914.86** | **5885.718** | **6035.568** | **0.279538** |
|  | 5 | -2892.87 | 5849.74 | 6020.998 | 0.242631 |
| Stunting<br>(HAZ< -2) | 1 | -839.848 | 1711.696 | 1802.497 | - |
|  | 2 | -650.679 | 1341.359 | 1454.861 | 0.336858 |
|  | 3 | -464.192 | 976.3846 | 1112.587 | 0.337546 |
|  | **4** | **-407.715** | **871.4293** | **1030.332** | **0.40443** |
|  | 5 | -344.902 | 753.8037 | 935.4063 |  |
| Underweight<br>(WAZ< -2) | 1 | -2376.03 | 4784.068 | 4873.619 | - |
|  | 2 | -2232.69 | 4505.378 | 4617.316 | 0.216863 |
|  | 3 | -2160.18 | 4368.352 | 4502.677 | 0.157158 |
|  | 4 | -2118.11 | 4292.216 | 4448.929 | 0.153133 |
|  | **5** | **-2120.83** | **4305.662** | **4484.763** | **0.43731** |

AIC- Akaike's information criterion; BIC- Bayesian information criterion
Number of optimal classes for each nutritional outcome is bolded.
\*Respective baseline anthropometric values Age, sex, wealth quintile, modified Vesikari score, and antibiotic use included in LCMM models

**Table S2.** Factors Associated with Worsening and Stagnating Post-diarrheal Nutritional Trajectories in the Wasting Cohort.

| Predictor | Category | Base Outcome (Improving) |  |  |  |  |  |
| --- | --- | --- | --- | --- | --- | --- | --- |
|  |  | Worsening |  |  | Stagnating |  |  |
|  |  | RR (95% CI) | p_value | aRR (95% CI) | RR (95% CI) | p_value | aRR (95% CI) |
| Age categories | 6-11m | Ref |  | Ref | Ref |  | Ref |
|  | 12-17m | 0.95 (0.48–1.86) | 0.874 | 0.88 (0.42–1.84) | 0.94 (0.67–1.31) | 0.712 | 0.90 (0.63–1.28) |
|  | 18-23m | **0.31 (0.11–0.92)** | **0.034** | **0.28 (0.09–0.86)** | 0.78 (0.54–1.13) | 0.183 | 0.76 (0.51–1.13) |
|  | 24-35m | **0.20 (0.05–0.87)** | **0.031** | 0.23 (0.05–1.02) | 0.68 (0.44–1.04) | 0.072 | 0.67 (0.41–1.07) |
| Sex | Male | 1.00 (0.54–1.85) | 0.996 | 0.92 (0.48–1.76) | 1.23 (0.93–1.61) | 0.141 | 1.23 (0.93–1.64) |
| No. of children <5 yrs in household | <3 | Ref |  | Ref | Ref |  | Ref |
|  | ≥3 | 0.76 (0.38–1.52) | 0.431 |  | 1.2 (0.91–1.59) | 0.202 |  |
| Final Quintile | Quintile 1- Least wealthy | Ref |  | Ref | Ref |  | Ref |
|  | Quintile 2 | **0.42 (0.19–0.95)** | **0.037** | 0.49 (0.21–1.15) | **0.67 (0.47–0.96)** | **0.029** | **0.64 (0.44–0.94)** |
|  | Quintile 3 | 0.51 (0.22–1.17) | 0.111 | 0.60 (0.25–1.47) | **0.68 (0.47–0.99)** | **0.049** | 0.74 (0.49–1.11) |
|  | Quintile 4 | 0.58 (0.23–1.47) | 0.249 | 0.70 (0.24–1.99) | **0.59 (0.37–0.93)** | **0.022** | 0.66 (0.41–1.08) |
|  | Quintile 5- Wealthiest | - | - | - | **0.46 (0.23–0.94)** | **0.034** | 0.56 (0.26–1.22) |
| Mother's education<sup>β</sup> | High Education | Ref |  | Ref | Ref |  | Ref |
|  | Low Education | **2.69 (1.19–6.1)** | **0.018** | 2.19 (0.89–5.37) | 1.21 (0.91–1.62) | 0.193 | 1.23 (0.87–1.72) |
| Father's education<sup>β</sup> | High Education | Ref |  | Ref | Ref |  | Ref |
|  | Low Education | 1.89 (0.94–3.81) | 0.075 | 1.27 (0.59–2.72) | 1.14 (0.87–1.51) | 0.343 | 1.02 (0.74–1.41) |
| Caregiver occupation<sup>γ</sup> | No employment | Ref |  | Ref | Ref |  | Ref |
|  | Formal employment | - | - | - | 1.29 (0.47–3.5) | 0.622 | 1.64 (0.58–4.67) |
|  | Informal employment | 0.68 (0.26–1.75) | 0.425 | 0.64 (0.23–1.76) | **0.65 (0.43–0.99)** | **0.043** | 0.68 (0.44–1.06) |
| Temperature |  | 1.27 (0.92–1.76) | 0.146 |  | 1.07 (0.91–1.25) | 0.4 |  |
| Respiratory rate* | Normal | Ref |  | Ref | Ref |  | Ref |
|  | Low | 0.75 (0.26–2.14) | 0.588 |  | 0.74 (0.46–1.19) | 0.22 |  |
|  | High | 0.32 (0.03–2.91) | 0.31 |  | 0.69 (0.33–1.45) | 0.326 |  |
| Heart rate* | Normal | Ref |  | Ref | Ref |  | Ref |
|  | Low | 0.44 (0.18– | 0.072 | 0.50 (0.19– | 1.37 (0.75–2.49) | 0.306 | 1.33 (0.71– |
|  |  | 1.08) |  | 1.35) |  |  | 2.50) |
|  | High | - | - | - | 1.03 (0.27–3.98) | 0.963 | 1.35 (0.31–5.78) |
| Stool Frequency |  | 0.99 (0.90–1.10) | 0.876 |  | 0.99 (0.95–1.04) | 0.661 |  |
| Diarrhea days |  | 1.09 (0.88–1.34) | 0.432 | 1.05 (0.83–1.33) | **1.11 (1.01–1.22)** | **0.028** | **1.11 (1.01–1.23)** |
| Dysentery | Yes | 0.60 (0.21–1.69) | 0.332 |  | 1.04 (0.72–1.51) | 0.833 |  |
| Vomiting | Yes | 1.16 (0.62–2.16) | 0.65 |  | 1.02 (0.78–1.35) | 0.865 |  |
| Vomit frequency |  | 0.99 (0.84–1.17) | 0.937 |  | 0.95 (0.88–1.03) | 0.208 |  |
| Vomit days |  | 1.19 (0.99–1.43) | 0.069 | 1.06 (0.84–1.35) | 0.98 (0.87–1.09) | 0.67 | 0.95 (0.82–1.11) |
| Fever | Yes | 1.15 (0.61–2.19) | 0.666 | 0.89 (0.44–1.81) | 0.82 (0.63–1.08) | 0.153 | 0.81 (0.60–1.09) |
| Lethargic | Yes | - | - |  | 0.49 (0.06–3.82) | 0.493 |  |
| Cough | Yes | 0.88 (0.45–1.70) | 0.695 |  | 0.88 (0.66–1.17) | 0.376 |  |
| Difficulty breathing | Yes | - | - |  | 0.37 (0.05–2.87) | 0.344 |  |
| Sunken eyes | Yes | 1.54 (0.76–3.11) | 0.227 |  | 1.23 (0.89–1.71) | 0.211 |  |
| Jaundice | Yes | - | - |  | 2.46 (0.45–13.48) | 0.301 |  |
| Drink or breastfeed | Normal/Not Thirsty | Ref |  | Ref | Ref |  | Ref |
|  | Drinks eagerly/Thirsty | 1.49 (0.76–2.9) | 0.245 |  | 0.95 (0.69–1.32) | 0.775 |  |
|  | Drinks poorly/Unable to drink | - | - |  | 1.46 (0.40–5.34) | 0.571 |  |
| Stridor | Yes | - | - |  | 2.45 (0.22–27.12) | 0.465 |  |
| Convulsion | Yes | - | - |  | 1.63 (0.17–15.75) | 0.672 |  |
| Oxygen saturation < 90% | Yes | - | - | - | 2.19 (0.67–7.17) | 0.195 | 1.93 (0.55–6.74) |
| Skin Turgor | Normal | Ref |  | Ref | Ref |  | Ref |
|  | Slowly | **2.67 (1.25–5.71)** | **0.012** | 1.87 (0.69–5.08) | **1.7 (1.14–2.52)** | **0.009** | **1.96 (1.18–3.27)** |
|  | Very slowly | - | - | - | 0.47 (0.06–3.66) | 0.472 | 0.38 (0.04–3.40) |
| Chest Indrawing | Yes | - | - |  | 0 (0–Inf) | 0.984 |  |
| Dehydration | None | Ref |  | Ref | Ref |  | Ref |
|  | Severe | - | - |  | 0.75 (0.22–2.56) | 0.648 |  |
|  | Some | 1.89 (0.97–3.69) | 0.062 |  | 1.16 (0.83–1.61) | 0.381 |  |
| Modified Vesikari Score | Mild | Ref |  | Ref | Ref |  | Ref |
|  | Moderate | 1.51 (0.65–3.52) | 0.341 | 1.13 (0.44–2.92) | 0.92 (0.63–1.34) | 0.671 | 0.90 (0.59–1.37) |
|  | Severe | **2.09 (1.05–4.15)** | **0.035** | 1.53 (0.60–3.90) | 1.03 (0.75–1.41) | 0.87 | 1.04 (0.66–1.64) |
| ORS | Yes | **2.08 (1.05–4.14)** | **0.036** | 1.30 (0.50–3.36) | 0.74 (0.49–1.1) | 0.136 | **0.59 (0.36–0.97)** |
| IV fluids | Yes | 0.69 (0.09–5.14) | 0.718 | 0.53 (0.07–4.26) | 1.75 (0.96–3.2) | 0.07 | 1.92 (0.94–3.95) |
| Zinc | Yes | 0.67 (0.16–2.8) | 0.58 |  | 0.71 (0.39–1.29) | 0.257 |  |
| Food supplement at enrolment | Yes | - | - |  | 1.37 (0.4–4.64) | 0.613 |  |
| Food supplement for home use | Yes | 2.00 (0.69–5.78) | 0.2 |  | 1.01 (0.55–1.86) | 0.977 |  |
| WHO-recommended antibiotic | Yes | 0.88 (0.47–1.64) | 0.696 | 0.72 (0.35–1.46) | **0.73 (0.56–0.96)** | **0.027** | **0.69 (0.51–0.94)** |
| Enrolment HAZ |  | 0.83 (0.66–1.05) | 0.126 | 0.79 (0.60–1.03) | 1 (0.9–1.11) | 0.99 | 1.00 (0.90–1.12) |
| Drinking water | Improved | Ref |  | Ref | Ref |  | Ref |
|  | Unimproved | 1.04 (0.14–7.79) | 0.972 |  | 0.67 (0.23–1.92) | 0.455 |  |
| Sanitation | Improved | Ref |  | Ref | Ref |  | Ref |
|  | Unimproved | 1.15 (0.48–2.77) | 0.76 |  | 0.84 (0.55–1.28) | 0.415 |  |
| Rotavirus age-appropriate vaccination | Yes | 0.84 (0.30–2.41) | 0.753 | 1.17 (0.37–3.64) | 0.73 (0.47–1.14) | 0.163 | 0.80 (0.50–1.29) |
| Exclusive Breast feeding | Yes | **0.31 (0.16–0.63)** | **0.001** | **0.37 (0.18–0.77)** | **0.73 (0.56–0.95)** | **0.022** | **0.73 (0.55–0.96)** |

RR- risk ratios; aRR- adjusted Risk ratios
<sup>b</sup> High education- some secondary, and secondary school or greater; Low education: No education, less than primary, primary school only, and Koranic school only.
<sup>\*</sup> No employment-Not employed, housewife, and student; Formal employment- professional; Informal employment-business (self-employed), business (other employer), casual laborer, and farmer.
<sup>\*</sup>Cutoffs for heart rate and respiratory rate based on the Pediatric Advanced Life Support (PALS) guidelines.
<sup>b</sup>Modified 20-point vesikari score

**Table S3.**
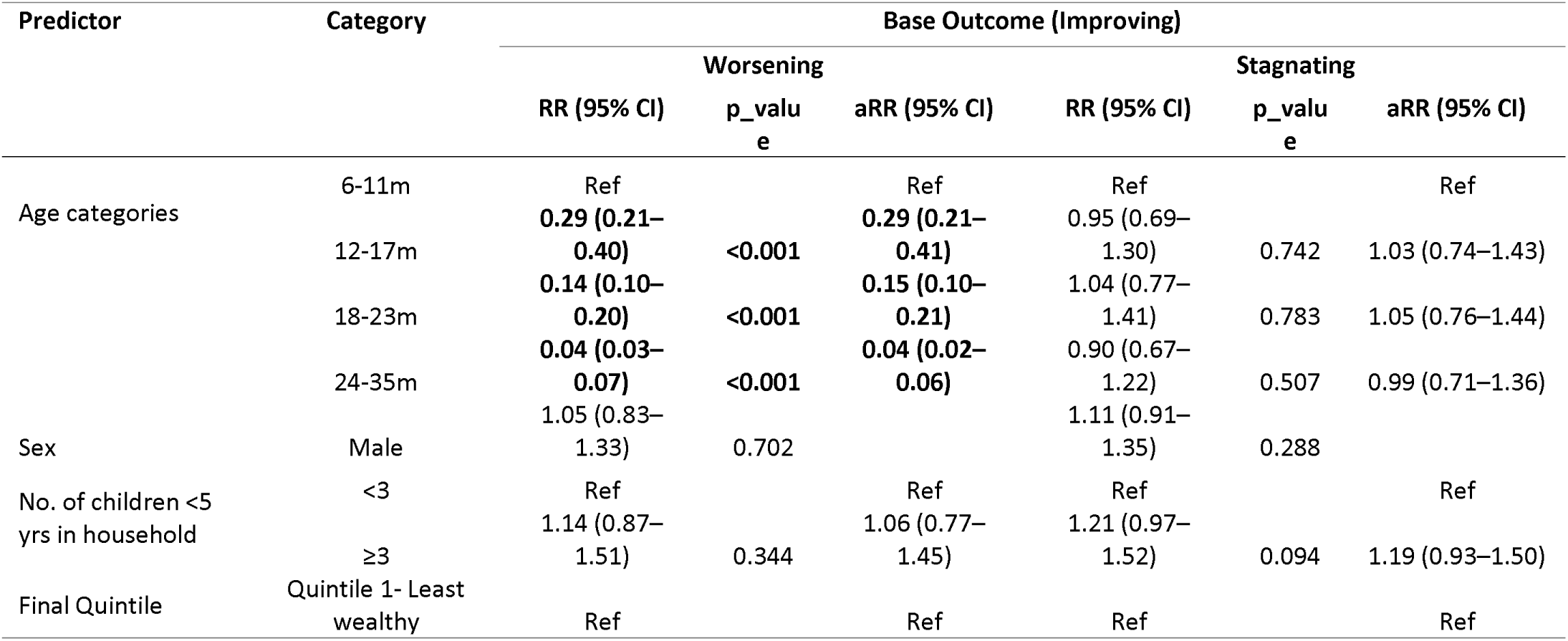
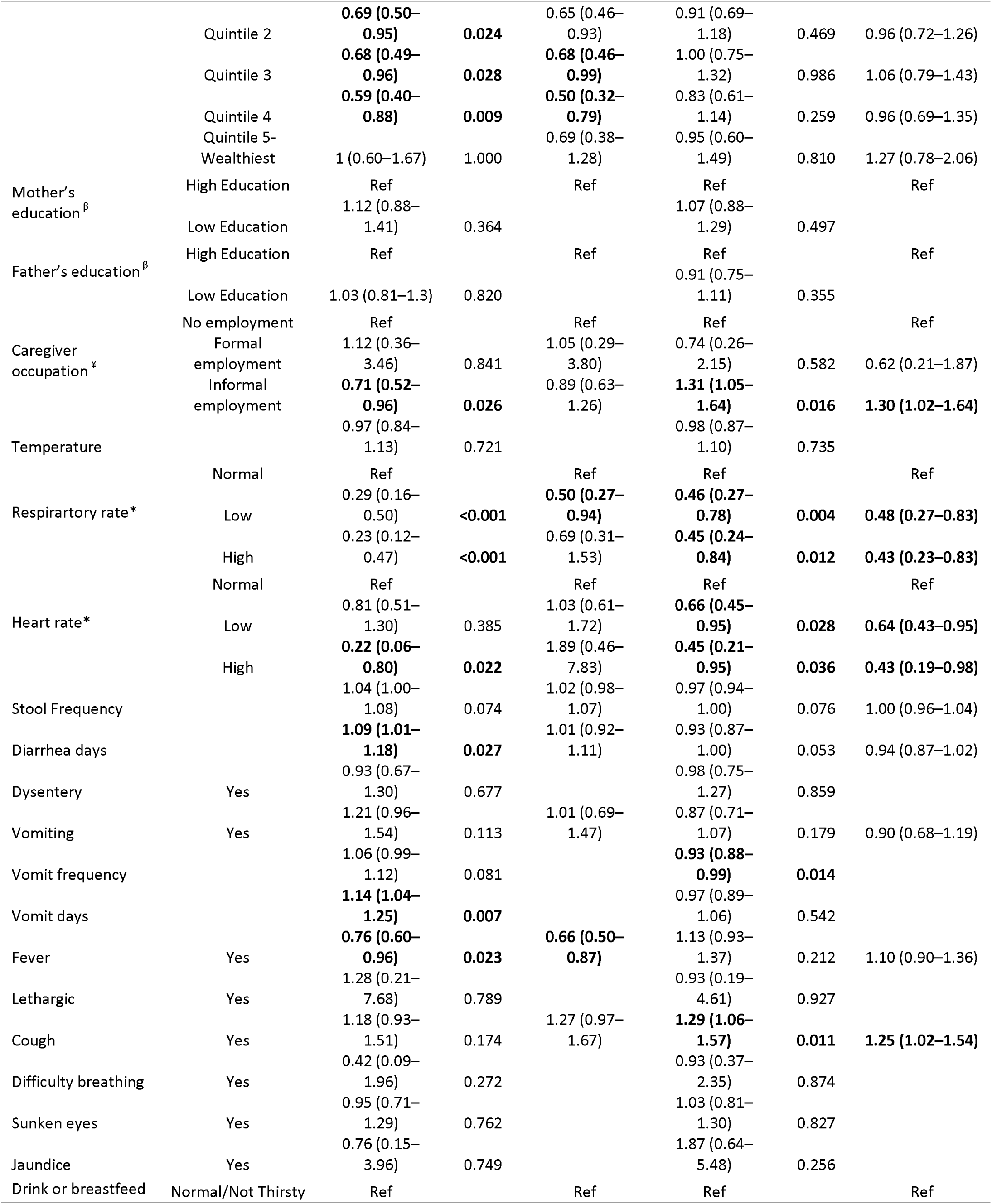
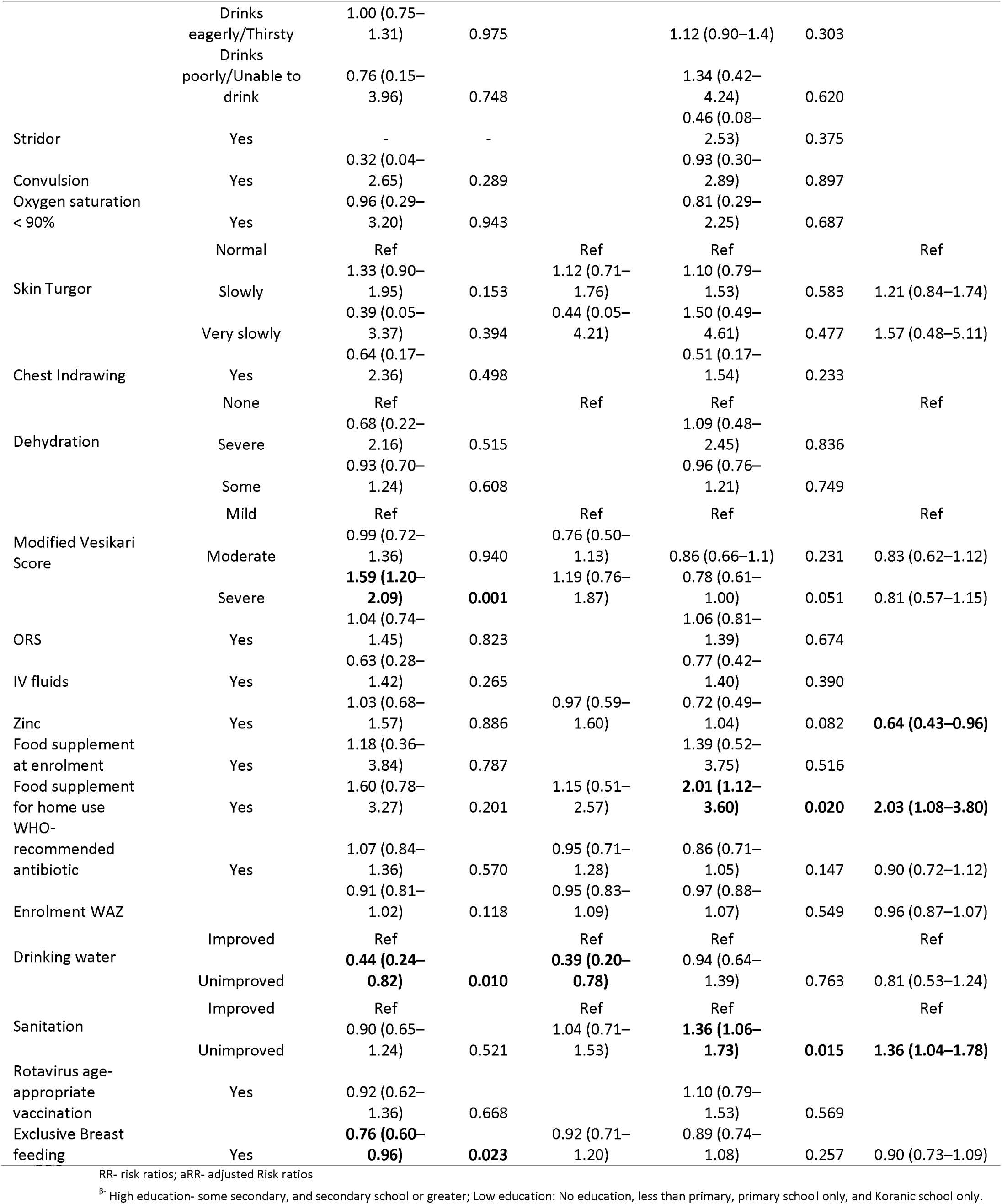
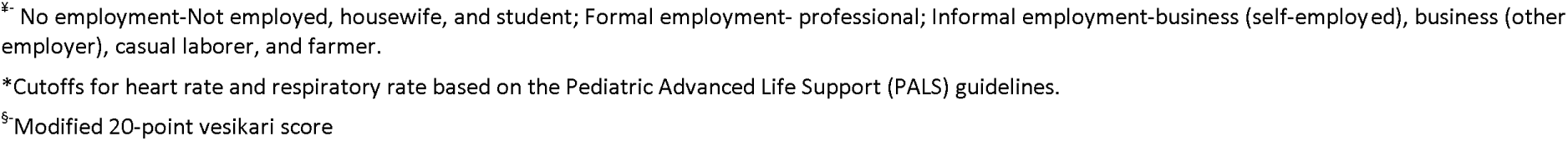
Factors Associated with Worsening and Stagnating Post-diarrheal Nutritional Trajectories in the Stunting Cohort.

**Table S4.**
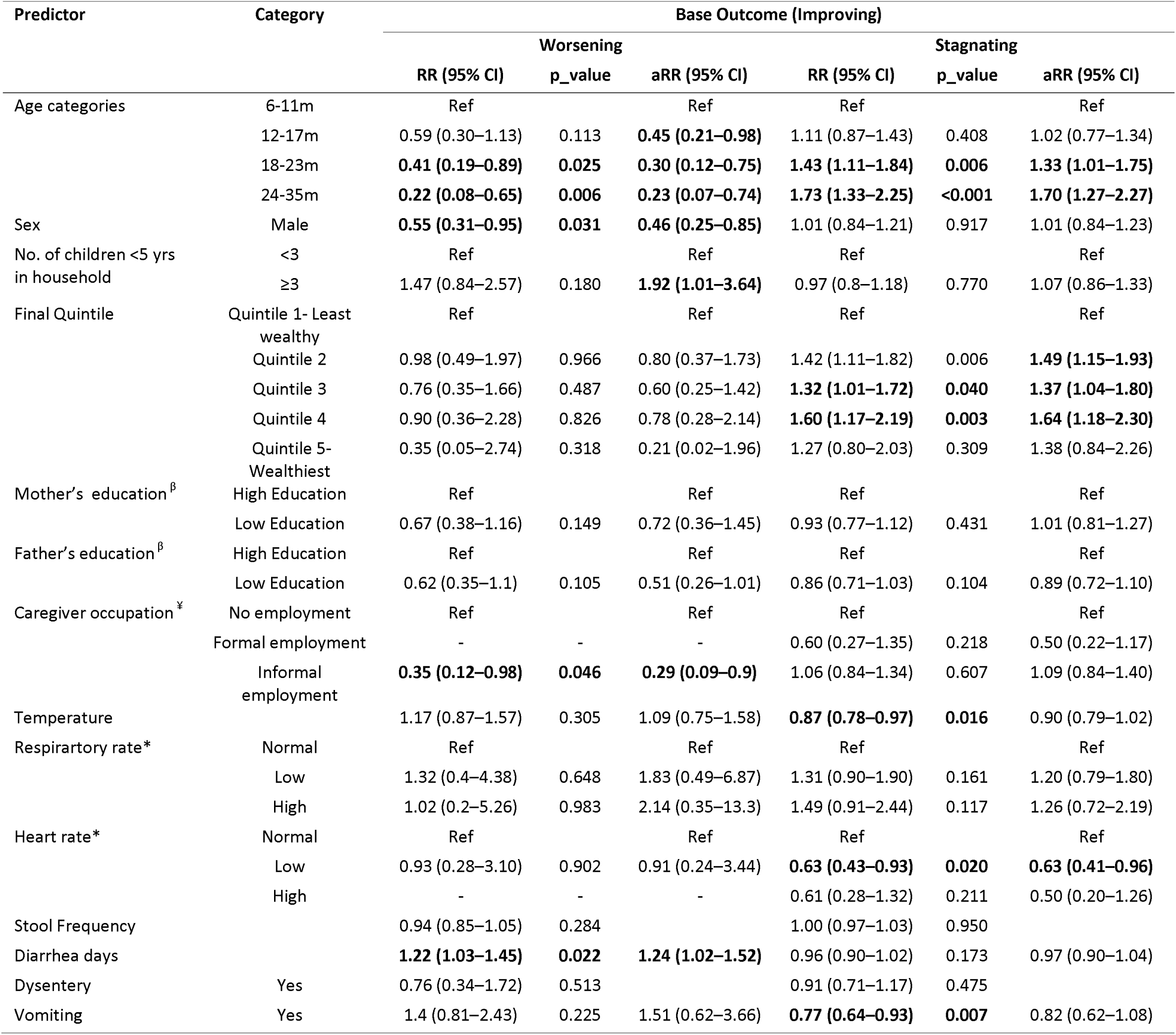
Factors Associated with Worsening and Stagnating Post-diarrheal Nutritional Trajectories in the Underweight Cohort.

